# Muscle proteins in plasma associate to distinguished phenotypes in amyotrophic lateral sclerosis

**DOI:** 10.64898/2026.07.14.26357727

**Authors:** Louisa Azizi, Inci S. Aksoylu, María Bueno Álvez, Juliette Foucher, Alexander Juto, Christina Seitz, Raymond Press, Kristin Samuelsson, Ulf Kläppe, Mathias Uhlén, Fredrik Edfors, Sofia Bergström, Fang Fang, Peter Nilsson, Linn Öijerstedt, Anna Månberg, Caroline Ingre

## Abstract

**Background:** Amyotrophic lateral sclerosis (ALS) is a neurodegenerative disease characterized by death of upper and lower motor neurons, usually presented with clinical heterogeneity. Fluid biomarker development remains dominated by neurofilament light chain (NEFL), a marker of neuroaxonal injury. NEFL is however unspecific to ALS and its phenotypes and there is currently a lack of biomarkers that capture ALS heterogeneity such as onset site and ALS-frontotemporal spectrum disorder (ALS-FTSD). Therefore, we investigated whether plasma proteomics could reveal pathway-level signatures that stratify and explain ALS heterogeneity.

**Methods:** We profiled ∼5,400 plasma proteins (Olink Explore HT) in 299 patients with ALS and 50 age- and sex comparable healthy controls. We used two complementary analytic frameworks: (i) differential protein abundance analysis to identify altered proteins in ALS and across clinical subgroups, and (ii) weighted gene correlation network analysis (WGCNA) to identify coordinated protein modules and relate them to ALS diagnosis and to ALS-specific clinical traits (site of onset, ALS-FTSD, ALS functional rating scale-revised (ALSFRS-R) score, and plasma NEFL).

**Results:** Differential abundance analysis identified 56 proteins altered in ALS versus controls, of which 40 were increased. WGCNA identified 11 co-expression modules, with ALS samples having the strongest correlation to a protein module (n=51) highly enriched for muscle-related proteins. Out of the 40 proteins that had increased expression levels, 29 overlapped with the muscle-enriched protein module, indicating that muscle related proteins are the dominant circulating proteomic signature in ALS. This signal extended to clinical stratification: spinal-onset patients showed a strong positive association with the muscle-module. Further, differential abundance analysis of spinal- versus bulbar-onset ALS identified changes that mapped predominantly to the same module, supporting a molecular signature of onset phenotype. In contrast, cognitive status (ALS-FTSD) mapped to distinct modules enriched for extracellular matrix/cell-adhesion pathways, consistent with a separable biological axis of disease heterogeneity. Although multiple modules correlated with NEFL, trait-specific signatures were not fully explained by neuroaxonal injury. Notably, the muscle-enriched module increased with higher NEFL and lower ALSFRS-R, supporting its interpretation as a severity-linked, muscle-involvement proxy.

**Conclusions:** Large-scale plasma proteomics reveals that heterogeneity in ALS reflects underlying biological structures. We identified a dominant muscle-associated protein network that distinguished ALS patients from controls and correlated with disease onset phenotype and severity, alongside distinct protein networks linked to ALS-FTSD. By integrating differential protein abundance with network-based analysis, we defined pathway-level biomarker signatures that extend beyond NEFL, enabling biologically informed patient stratification and improved therapeutic monitoring.

## Introduction

Amyotrophic lateral sclerosis (ALS) is a progressive neurodegenerative disease defined by degeneration of upper and lower motor neurons and a median survival of approximately 2–4 years from symptom onset.^1,2^ The clinical presentation varies between individuals, with substantial differences in progression rate, survival, site of onset and cognitive involvement, indicating biological and clinical heterogeneity.^1,2^ Cognitive and behavioral impairment within the ALS–frontotemporal spectrum disorder (ALS-FTSD) affects up to 30–50% of patients and represents another major source of phenotypic variability.^3–6^

Therapeutic options in ALS remain limited, reflecting incomplete understanding of the underlying pathophysiology, and there is currently no established biomarker that capture disease activity and diversity. Multiple disease processes, including neuronal hyperexcitation, toxic protein accumulation, mitochondrial dysfunction, and neuroinflammation have been implicated, yet their relative contributions differ across patients and disease stages.^4,7,8^ This heterogeneity complicates not only diagnosis but also clinical trial design, where biologically distinct subgroups may respond differently to targeted interventions and have different biomarkers are used to evaluate the response.

ALS diagnosis continues to rely primarily on clinical examination and electromyography, with supportive laboratory testing, rather than molecular markers with high specificty.^9^ Consequently, diagnostic delay remains substantial, with population-based studies reporting median delays of 10–12 months from symptom onset, limiting timely interventions and trial enrollment.^9,10^ Fluid biomarkers capable of detecting early disease and monitoring progression are therefore critical.

Neurofilament light chain (NEFL) in cerebrospinal fluid (CSF) and blood has emerged as the most reliable biomarker of neuroaxonal damage in ALS, consistently correlating with survival.^11–14^ However, while NEFL provides strong prognostic value and aids diagnostic discrimination from mimics, it is not specific for ALS and is increased in several neurological disorders.^15,16^

Additional candidates, including and muscle-derived proteins such as troponin T, suggest that peripheral and neuromuscular remodeling may represent complementary disease axes not captured by neuroaxonal markers alone.^17–21^ Emerging proteomic approaches demonstrate that circulating protein signatures can predict ALS diagnosis, applicable for future diagnostics and early interventions.^22,23^ However, there is a lack of proteomic signatures that reflect ALS heterogeneity and the different pathway-level mechanisms amongst these phenotypes. Such biomarker stratification is critical to support mechanism-based therapeutic development, as well as patient selection and monitoring in clinical trials. The aim of this study was therefore to characterize phenotypic heterogeneity in ALS by using network-based plasma proteomics and identify associated protein signatures, to improve understanding of disease mechanisms and biomarkers explaining these.

## Materials and method

### Study materials description

The study population was derived from the ALSrisc Study, a case-control study conducted since 2016 at the ALS Clinical Research Centre at Karolinska Institutet in Stockholm, Sweden.^24^ Patients with a newly diagnosed ALS according to the Gold Coast diagnostic criteria were consecutively recruited. Siblings and spouses to the patients were recruited as controls.^25^ A total of 349 individuals were included in the present analysis, including 299 patients with ALS and 50 controls (19 siblings, 31 spouses). Clinical variables included age at plasma sampling and sex for all subjects, and site of symptom onset, cognitive status, body mass index (BMI), genetic status, and amyotrophic lateral sclerosis functional rating scale revised (ALSFRS-R) scores for patients.

Site of symptom onset was classified as the location of the first motoric symptom, categorized as spinal (including respiratory and trunk) or bulbar. Whole-genome sequencing was performed in 239 patients, with bioinformatic analysis restricted to a predefined panel with 44 associated with ALS.^26^ Absence of pathogenic variants within this panel was classified as a negative genetic screen.

Survival was defined as the date of tracheostomy or death, whichever occurred first. The last date of follow-up was October 29^th^, 2025. Patients with survival time < 1.5 years from symptom onset were considered short survivors, ≤1.5 - ≤5.5 years as intermediate survivors, and > 5.5 years as long survivors, thresholds adapted to reflect the distribution of survival times in our cohort.^27^ In cases when the patients were still alive at the end of follow-up, they were categorized as long survivors if the survival time had exceeded 5.5 years on that date, otherwise they were considered alive and not eligible to be categorized (N=21).

Disease functional impairment and progression rate were assessed using the ALSFRS-R score. The ALSFRS-R is a validated 12-item scale ranging from 48 (normal function) to 0 (maximal disability), covering bulbar, fine motor, gross motor, and respiratory functional domains.^28^ Only ALSFRS-R scores collected within 3 months of plasma sampling were included (missing N=48). Delta ALSFRS-R (Δ ALSFRS-R) was calculated by dividing the difference between a patient’s first and last recorded ALSFRS-R score by the time interval in months and used as an indicator of functional decline rate over the disease course. Slow progressors were defined as a decline of <0.5 ALSFRS-R points per month, intermediate progressors as a decline of ≥0.5 and <1.5 points per month, and fast progressors as a decline of ≥1.5 points per month, thresholds set to capture extreme phenotypes.

Cognitive status (ALS-FTSD or non-FTSD-ALS) was defined by either a total score below 108 points on the Swedish-validated Edinburgh Cognitive and Behavioral ALS Screen (ECAS) or a clinical diagnosis of frontotemporal dementia (FTD).^29,30^ FTD was established by a specialist at a tertiary clinic following comprehensive evaluation, including neuroimaging and comprehensive neuropsychological testing. Patients diagnosed with FTD at the time of plasma sampling or within six months thereafter were categorized as ALS-FTSD. ECAS screening within one month before or up to six months after plasma sampling was included.

### Sample collection and proteome profiling

Venous blood samples were collected in EDTA tubes within a median of 36 days from diagnosis date and stored in -80°C at Karolinska Institutet Biobank. Proteomics profiling was performed using the Olink Explore HT proteomics platform at the SciLifeLab Affinity Proteomics and National Genomics Infrastructure (NGI) in Uppsala, as part of the Human Disease Blood Resource within the Human Protein Atlas (www.proteinatlas.org).^31^

Olink Explore HT is a high-throughput proteomics platform based on Proximity Extension Assay (PEA) technology, enabling the simultaneous measurement of >5400 protein assays. In short, target proteins are recognized by pairs of antibodies, each conjugated to complementary, assay-specific oligonucleotides. When both antibodies bind the same protein, their oligonucleotides come into proximity, allowing hybridization and enzymatic extension to form a unique DNA sequence, ensuring specificity of protein detected. The resulting unique DNA barcodes are amplified and quantified by next-generation sequencing.

Normalized Protein eXpression (NPX) values were generated using the NPX map software by using internal control samples to adjust for technical variation across samples and plates. Quality control (QC) procedures included controls for incubation, extension, detection and inter-plate reference samples. Measurements failing predefined quality thresholds were flagged. After quality control, data was log2-transformed to generate NPX values and later normalized following the intensity normalization procedure to minimize technical variation across samples and plates. NPX represents relative protein abundance and enables comparison of the same protein across samples.

Additional data filtering was applied to ensure high-quality measurements. All data points failing quality control were removed prior to downstream analyses. Samples with >50% of measurements flagged for QC failure were excluded. Furthermore, proteins from dilution block 8 (n = 68, dilution 1:100,000), were excluded due to technical issues identified by the provider. For downstream analyses, proteins were excluded if detectable below the assay-specific limit of detection (LOD) in >85% of all samples which underwent analysis; ALS-samples (n=299), controls (n=50) and ALS mimics (n=84). The last group was excluded in the rest of the study. A total of 2412 proteins were included in the down stream analysis.

### Bioinformatics and statistical analysis

NPX values were adjusted for global protein levels (defined as median NPX), age, and sex using regression-based adjustment with removeBatchEffect function, to account for confounders, due to global variance in protein levels (eFigure 1 in Supplement 1). Analyses were performed using the limma-package (v3.66).^32^

### Differential Protein Abundance Analysis

Differential abundance was then assessed using linear models with Empirical Bayes moderation via the limma package (v3.66).^32^ Three group-wise comparisons were performed: (i) ALS vs. controls; (ii) ALS-FTSD vs. non-FTSD ALS, with additional adjustment for ALSFRS-R, BMI, and site of symptom onset; and (iii) Spinal vs. Bulbar onset, adjusted for ALSFRS-R and BMI. Proteins were defined as significant based on an absolute-fold change > 0.5 and an FDR-adjusted p-value < 0.05. Results were visualized with volcano plots, color-coded according to weighted gene correlation network analysis (WGCNA) module assignment, described below.

### Weighted gene correlation network analysis

WGCNA was applied to detect modules with highly correlated proteins using normalized protein abundance data in samples from ALS patients and controls using the WGCNA package (v1.73).^33^ In WGCNA, NEFL was excluded in the creation of modules so it could be retained as an independent variable in the module trait association analysis described below. Soft-thresholding power was identified with the WGCNA packages’ pickSoftThreshold function and selected based on scale-free topology (R² >0.8), low mean connectivity, and biologically interpretable module eigengene (ME) sizes, resulting in a soft threshold of 5 (eFigure 2-3 in Supplement 2). Network construction was performed using topological overlap matrix (TOM) dissimilarity, with the Tomsimilarity function. Modules were identified using hierarchical clustering with dynamic tree cutting (deepSplit = 2) and a minimum module size of 30 proteins. Hierarchical clustering was performed using hclust in flashClust package (v.1-01-2) (distance metric=1-TOM, linkage method=average).^34^ The gene dendrogram was created with plotDendroAndColors in WGNCA-package (hang = 0.03, guidehang= 0.05) (eFigure 4 in Supplement 1). Proteins that did not meet criteria for module membership were assigned to the grey module, which does not represent a functional correlation module and was excluded from downstream analysis. Module merging was evaluated using a cut height of 0.15 (eFigure 5 in Supplement 1). No modules met criteria for merging.

Module-trait association analyses were conducted using two models: one including comparison between ALS and controls, and one including comparisons within the ALS group including patients with different clinical traits, based on the same set of MEs. Correlations were assessed using Pearsons’s correlation and significance was defined as p-value<0.05. For each ME, hub proteins were defined by module membership (MM) > 0.8, calculated using Pearsons’s correlation for intramodular connectivity, and the top 10 proteins per module were selected in descending order of MM. All protein modules were subjected to Gene Ontology (GO) biological process enrichment analysis with all proteins passing LOD as background and annotation were obtain from org.Hs.eg.db.^35^ The associations between these hub proteins and clinical outcomes were subsequently modeled using logistic regression via the glm.fit function (stats package v.4.5.1). A logit link function was used, expressed as:

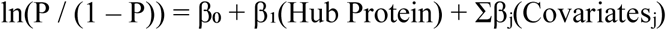

In the analysis comparing ALS to controls, no additional covariates were included beyond the initial prior adjustments (adjusted for age, sex, and median NPX). When comparing site of symptom onset (spinal vs. bulbar), ALSFRS-R score and BMI were included as covariates, while models for ALS-FTSD vs. non-FTSD-ALS were adjusted for site of onset, ALSFRS-R score, and BMI. For these logistic models, proteins were deemed relevant if FDR-adjusted p-value was < 0.05 and area under the curve (AUC) was > 0.65.

To assess associations between hub protein levels and ALSFRS-R, ECAS, and plasma NEFL levels, robust linear regression was employed, defined by the model:

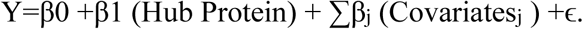

In these models, Y represented the ALSFRS-R score (adjusted for site of onset and BMI), ECAS score (adjusted for site of onset, ALSFRS-R score, and BMI), or plasma NEFL levels (adjusted for site of onset, ALSFRS-R score, and BMI).

All data pre-processing, statistical analysis, and illustrations were performed in R version 4.5.1.^36^ Benjamin-Hochberg was applied to adjust p-values for multiple comparison, with an FDR threshold < 0.05 used in all analysis except for module trait association analyses.

## Results

### Study population characteristics

This study included 299 ALS patients and 50 controls. Clinical and demographic characteristics of the study population are summarized in Table 1. The cohort was representative of ALS population, described in large multicenter cohort studies, in terms of site of symptom onset (61.2% with spinal onset and 38.1% with bulbar onset), ALS-FTSD status (23.7% with ALS-FTSD), and genetic status (15.9% with a mutation).^1,2^ The median ALSFRS-R score at the time of plasma sampling was 39. Based on the rate of functional decline (Δ ALSFRS-R), 25.1% were classified as fast progressors, 42.8% as intermediate progressors, and 14.7% as slow progressors. A majority of patients were classified as intermediate survivors (62.2%), while smaller proportions were categorized as long (16.4%) or short survivors (14.4%).

**Table 1.** Demographic and clinical characteristics of ALS patients and controls included in the study.

| **Characteristics** | **ALS patients**<br>**(n=299)** | **Controls**<br>**(n=50)** |
| --- | --- | --- |
| Sex, n (%) |  |  |
| Female | 131 (43.8) | 25 (50) |
| Age, median (IQR) | 67 (60–74) | 62.5 (56–69) |
| BMI, median (IQR) | 22 (6.8–24.6) |  |
| Onset site, n (%) |  |  |
| Bulbar | 115 (38.1) |  |
| Spinal | 184 (61.2) |  |
| ALSFRS-R at plasma sampling, median (IQR) | 39 (34–42) |  |
| $\Delta$ ALSFRS-R, n (%) | | |
| Fast | 75 (25.1) |  |
| Intermediate | 128 (42.8) |  |
| Slow | 44 (14.7) |  |
| Not categorized | 52 (17.4) |  |
| Survival category, n (%) |  |  |
| Long | 49 (16.4) |  |
| Intermediate | 186 (62.2) |  |
| Short | 43 (14.4) |  |
| Genetics, n (%) |  |  |
| Neg screen | 201 (67.2) |  |
| C9orf72 | 29 (9.7) |  |
| SOD1 | 5 (1.7) |  |
| OPTN | 1 (0.3) |  |
| TBK1 | 1 (0.3) |  |
| Not screened | 60 (20.1) |  |
| ECAS score at diagnosis,<br>median (IQR) | 112 (100–118) |  |
| ALS-FTSD status, n (%) |  |  |
| ALS-FTSD | 71 (23.7) |  |
| Non-ALS-FTSD | 90 (30.1) |  |
| Not categorized | 138 (46.2) |  |

ALS: amyotrophic lateral sclerosis; ALSFRS-R: ALS Functional Rating Scale-Revised; ALS-FTSD: ALS-frontotemporal spectrum disorder; BMI: body mass index; C9orf72: chromosome 9 open reading frame 72; IQR: interquartile range; OPTN: optineurin; SOD1: superoxide dismutase 1; TBK1: TANK-binding kinase 1.

The median ECAS score at diagnosis was 112 (IQR 100–118). ALS-FTSD was present in 23.7% of patients, 30.1% were classified as non-ALS-FTSD, and 46.2% could not be categorized.

### Plasma proteins differ in abundance between ALS patients and controls

To decipher ALS associated alterations of plasma proteins, we compared median NPX-, age- and sex-adjusted levels of 2412 proteins between patients and controls using limma based linear regression. This revealed 56 proteins with significantly altered levels, with 40 increased- and 16 decreased (Figure 1A, eTable 1 in Supplement 1). NEFL demonstrated the largest fold change and strongest statistical significance, consistent with its role as a marker of neuroaxonal damage. In contrast, ANT3 (antithrombin III) was among the most significantly decreased proteins, indicating a marked reduction in ALS compared with healthy controls (Figure 1A).

**Figure 1.**
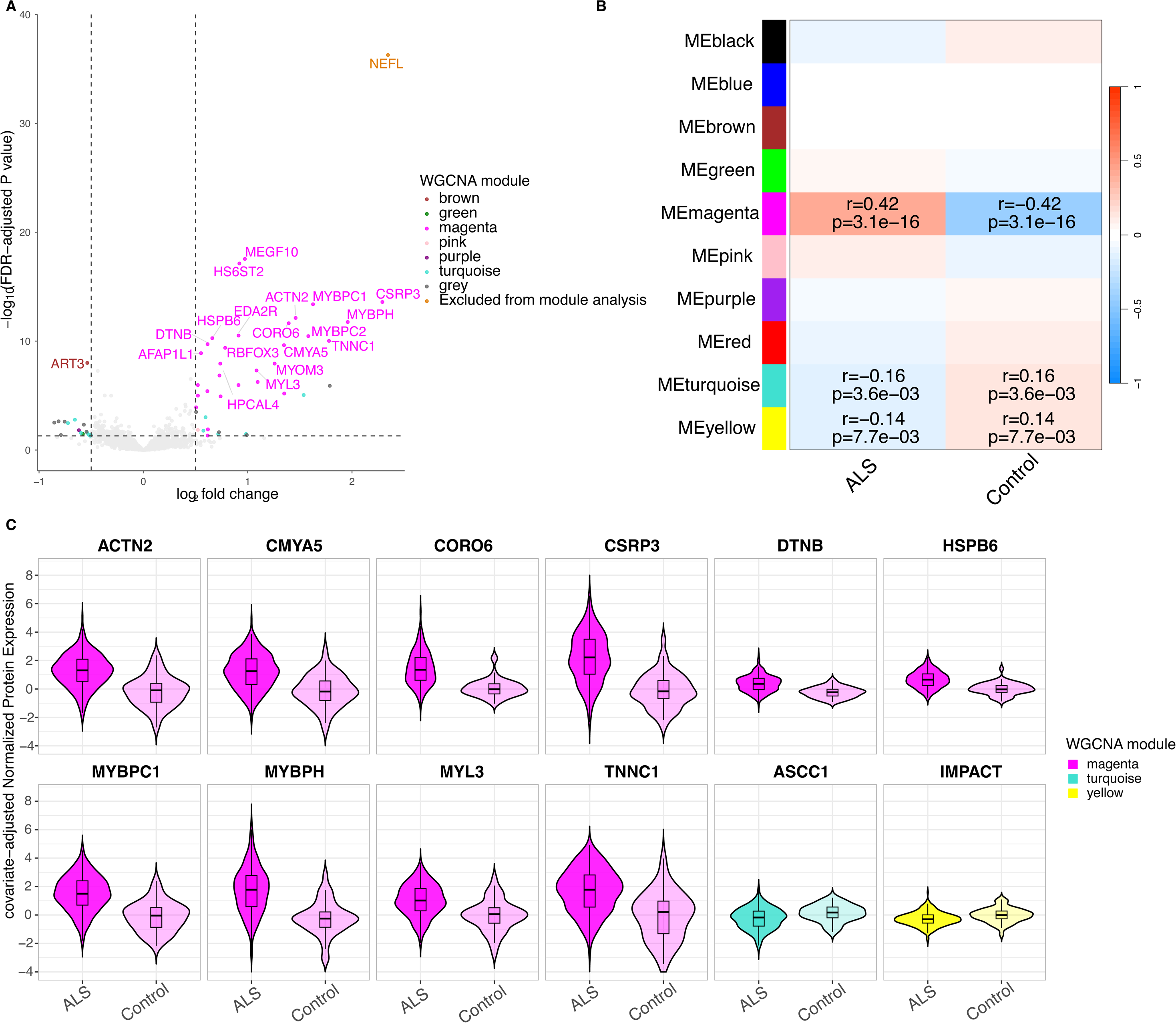
Proteomic analysis of ALS patients (n=299) versus controls (n=50). **A.** Volcano plot of differentially expressed proteins between ALS patients and controls. Dashed lines indicate inclusion thresholds (horizontal FDR adjusted p-value< 0.05; vertical |log₂-fold change| > 0.5). Total of 56 proteins met the inclusion criteria, including 40 increased- and 16 decreased levels in ALS patients compared with controls. The 20 most significant proteins are labeled, determined by FDR adjusted p-value. **B.** Module-trait association heatmap from weighted gene correlation network analysis (WGCNA). Modules (number of proteins): black (60), blue (184), brown (136), green (96), magenta (51), pink (57), purple (37), red (92), turquoise (1,012), and yellow (133). Color scale reflects Pearson correlation (r; red = 1, blue = −1). Correlation coefficients and p-values are displayed only for modules reaching statistical significance (p < 0.05). **C** Violin plots of covariate-adjusted (median normalized protein expression, age and sex) normalized protein expression values for the 12 hub proteins significantly associated with ALS versus controls.

### Correlated protein modules capture distinct biological processes in ALS

To characterize the biological processes underlying plasma proteome alterations in ALS, we applied WGCNA to identify correlated protein modules among the 2412 proteins. Our analysis identified 11 modules each summarized by module eigengenes, a numerical value that represent profiles of all proteins that are included in each module (eFigure 4 in Supplement 1). Modules identified were turquoise (1012 proteins), grey (533 proteins), blue (184 proteins), brown (136 proteins), yellow (133 proteins), green (96 proteins), red (92 proteins), black (60 proteins), pink (57 proteins), magenta (51 proteins) and purple (37 proteins) (eTable 2 in Supplement 1). All modules had 10 hub proteins that passed the defined thresholds (MM > 0.8), except for the purple, brown, and pink modules, which had 5, 6, and 9 hub proteins respectively (eTable 3 in Supplement 1). The results from GO-analysis showed that our modules were related to biological pathways including, but not limited to, muscle organization and function, synaptic signaling, extracellular matrix organization and neurodevelopment (Figure 2, Supplement 2). Three of the identified modules, namely magenta, turquoise, and yellow, included proteins significantly associated with having an ALS diagnosis when compared to controls (Figure 1B, eTable 4 in Supplement 1). The strongest positive correlation to ALS was observed for the magenta module (r = 0.42, p= 3 x 10^-16^), consisting of 51 proteins that were enriched for, but not limited to, muscle organization and contractile function (Figure 1B, Figure 2, eTable 4 in Supplement 1). On the other hand, we observed that ALS had negative correlations with immune-enriched turquoise (r=-0.16, p=0.004) and neurodevelopment-enriched yellow (r=-0.14, p=0.008) modules (Figure 1B, eTable 4 in Supplement 1). Notably, 72.5% (29 out of 40) of the proteins increased in ALS compared to controls were assigned to the muscle-enrichment module (Figure 1A).

**Figure 2.**
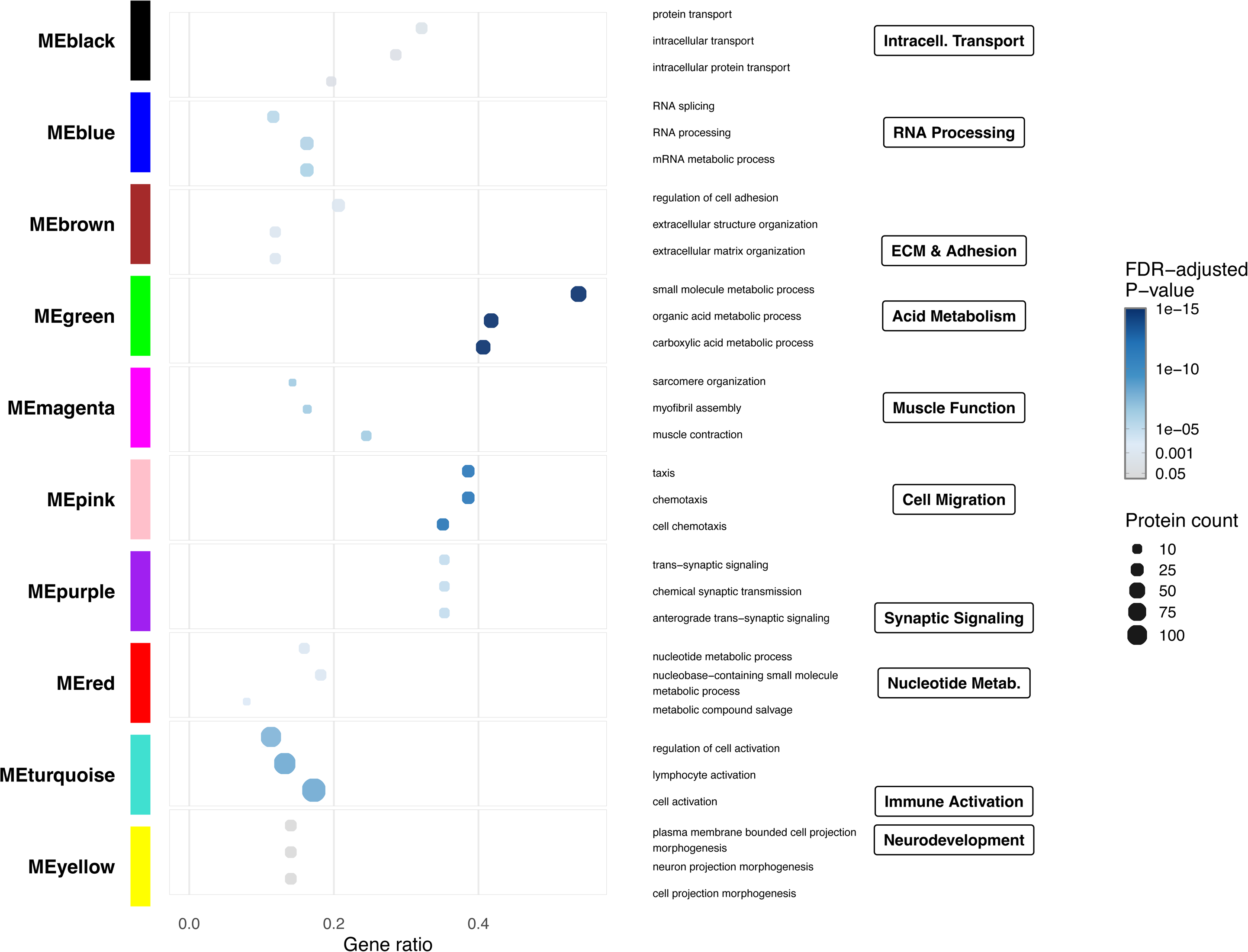
Functional enrichment analysis of correlation modules. Dot plot visualizing Gene Ontology (GO) terms enriched within identified gene modules (MEblack through MEyellow). The x-axis represents the gene ratio. Point size corresponds to protein count, while color intensity indicates the FDR corrected p-value. Summarized biological themes, such as synaptic signaling, immune activation, and neurodevelopment, are highlighted in a framed box for each module. Abbreviations: Intracell. Transport: intracellular transportation; ECM & Adhesion: extracellular matrix & adhesion; Nucleotide Metab: Nucleotide metabolism.

We next sought to link the identified module hub proteins to ALS diagnosis using logistic regression. A total of 12 hub proteins from muscle-enriched magenta, neurodevelopment-enriched yellow and immune-enriched turquoise were associated with ALS (Table 2, eTable 5 in Supplement 1). Our analysis showed that all hub proteins within the muscle-enriched magenta module were associated with ALS: MYBPC1, CSRP3, ACTN2, MYBPH, CORO6, HSPB6, DTNB, TNNC1, CMYA5, and MYL3 (Table 2, Figure 2). Increased plasma levels of these proteins were associated with increased odds of ALS and were higher in patients with ALS compared to controls (Table 2, Figure 1C). In contrast, for the neurodevelopment-enriched yellow and immune-enriched turquoise module, two of ten hub proteins were found with lower levels in ALS, namely IMPACT and ASCC1 respectively.

**Table 2.**
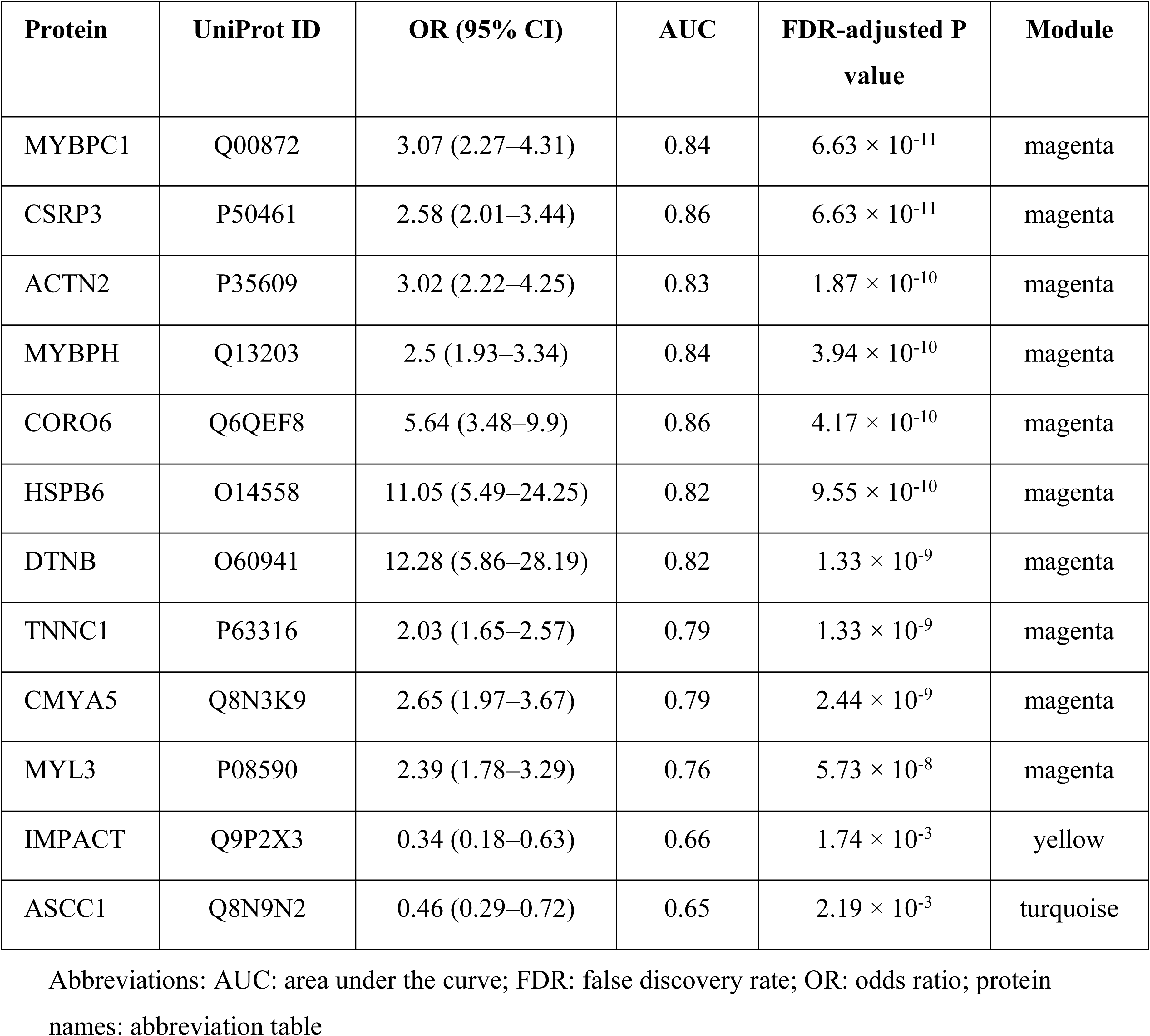
Logistic regression comparing levels of hub proteins in ALS patients (n=299) with controls (n=50). Only statistically significant proteins, defined as AUC > 0.65 and FDR adjusted p-value < 0.05, are included.

| Protein | UniProt ID | OR (95% CI) | AUC | FDR-adjusted P value | Module |
| --- | --- | --- | --- | --- | --- |
| MYBPC1 | Q00872 | 3.07 (2.27–4.31) | 0.84 | $6.63 \times 10^{-11}$ | magenta |
| CSRP3 | P50461 | 2.58 (2.01–3.44) | 0.86 | $6.63 \times 10^{-11}$ | magenta |
| ACTN2 | P35609 | 3.02 (2.22–4.25) | 0.83 | $1.87 \times 10^{-10}$ | magenta |
| MYBPH | Q13203 | 2.5 (1.93–3.34) | 0.84 | $3.94 \times 10^{-10}$ | magenta |
| CORO6 | Q6QEF8 | 5.64 (3.48–9.9) | 0.86 | $4.17 \times 10^{-10}$ | magenta |
| HSPB6 | O14558 | 11.05 (5.49–24.25) | 0.82 | $9.55 \times 10^{-10}$ | magenta |
| DTNB | O60941 | 12.28 (5.86–28.19) | 0.82 | $1.33 \times 10^{-9}$ | magenta |
| TNNC1 | P63316 | 2.03 (1.65–2.57) | 0.79 | $1.33 \times 10^{-9}$ | magenta |
| CMYA5 | Q8N3K9 | 2.65 (1.97–3.67) | 0.79 | $2.44 \times 10^{-9}$ | magenta |
| MYL3 | P08590 | 2.39 (1.78–3.29) | 0.76 | $5.73 \times 10^{-8}$ | magenta |
| IMPACT | Q9P2X3 | 0.34 (0.18–0.63) | 0.66 | $1.74 \times 10^{-3}$ | yellow |
| ASCC1 | Q8N9N2 | 0.46 (0.29–0.72) | 0.65 | $2.19 \times 10^{-3}$ | turquoise |

Abbreviations: AUC: area under the curve; FDR: false discovery rate; OR: odds ratio; protein names: abbreviation table

### Protein patterns are linked to ALS-specific clinical traits

Within the ALS patient group, we further explored associations between protein levels and specific clinical traits using logistic (site of symptom onset, cognitive status) or robust linear regression (NEFL levels and ALSFRS-R score).

### Muscle protein levels in plasma associate with spinal onset

Nine proteins were increased in patients with spinal compared to bulbar onset while no proteins were found to be decreased (Figure 3A, eTable 6 in Supplement 1). The muscle-enriched magenta module, which positively correlated with ALS diagnosis, was also positively correlated with spinal onset (r=0.3, p=1 x 10^-14^) (Figure 3B, eTable 7 in Supplement 1). Furthermore, we found that eight out of nine differentially abundant proteins were also a part of muscle-enriched magenta module indicating that muscle-function related proteins were enriched in spinal-onset patients (eTable 6 in Supplement 1). No other modules were significantly associated with site of symptom onset.

**Figure 3.**
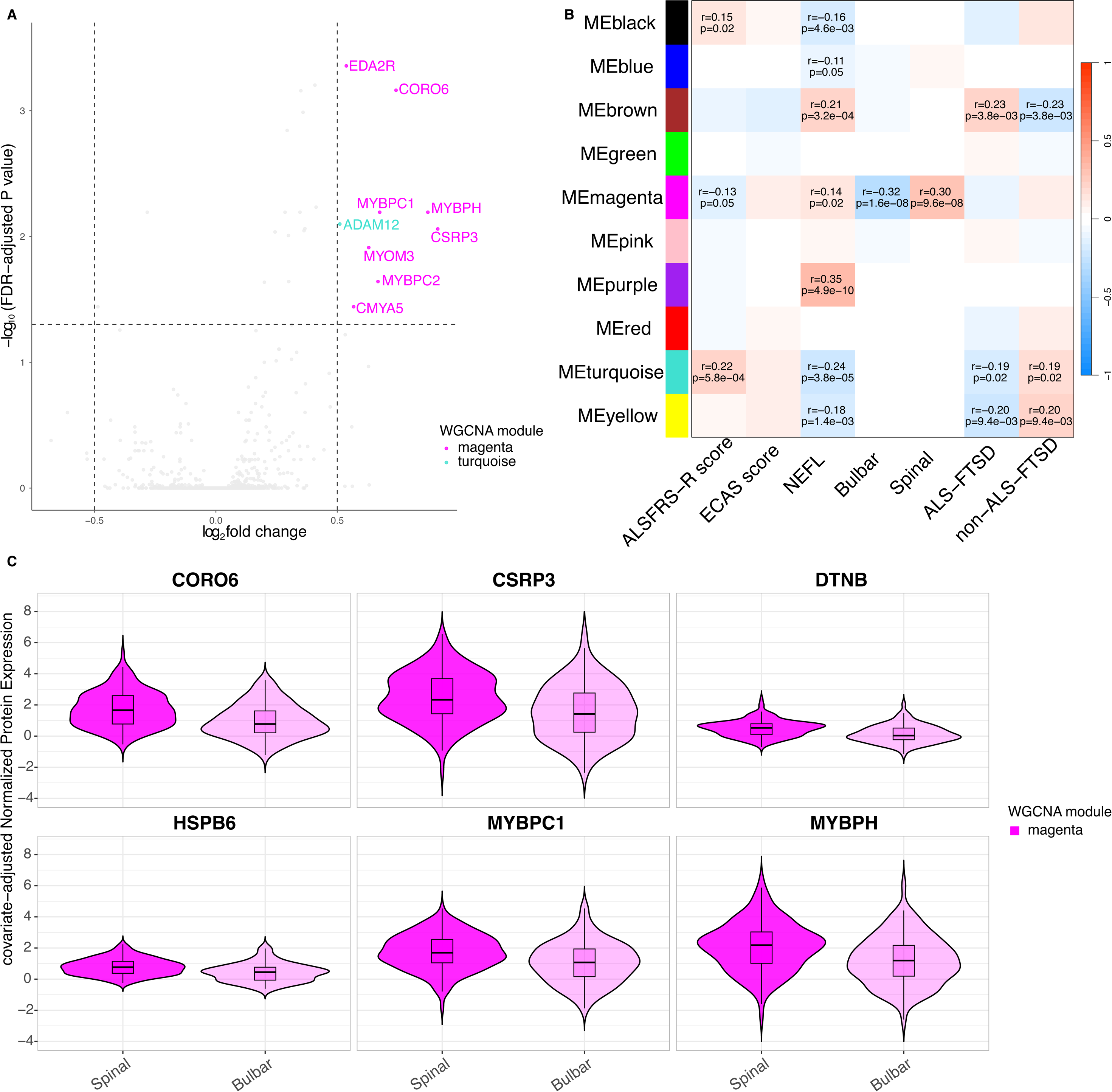
Proteomic analysis of ALS-specific clinical traits. **A**. Volcano plot of differentially expressed proteins between spinal (n=184) and bulbar (n=115) onset, adjusted for ALSFRS-R score and BMI. **B.** Module-trait association heatmap (WGCNA) for ALS-specific traits: ECAS score (n=156), ALSFRS-R score (n=251), spinal onset (n=184), bulbar onset (n=115), ALS-FTSD (n=71), and non-ALS-FTSD (n=90). Color scale reflects Pearson correlation (r; red = 1, blue = −1). Correlation coefficients and p-values are displayed only for modules reaching statistical significance (p < 0.05). **C.** Covariate-adjusted (median NPX, age and sex) normalized expression values of the 6 hub proteins significantly associated with spinal (n=184) compared to bulbar (n=115) onset, adjusted for ALSFRS-R score and BMI.

Following this, we assessed whether any specific proteins within the muscle-enriched magenta module were associated with spinal onset. Using logistic regression, we identified DTNB (OR 3.66, 95% CI 2.14–6.51, *p* = 2.22 x 10^-5^), CORO6 (OR 1.87, 95% CI 1.45 - 2.46, *p*= 2.22 x 10^-5^), HSPB6 (OR 3.24, 95% CI 1.97 – 5.5, *p*= 2.26 x 10^-5^), MYBPC1 (OR 1.6, 95% CI 1.28 – 2.02, *p*= 1.18 x 10^-4^), MYBPH (OR 1.45, 95% CI 1.21 – 1.74, *p*= 1.18 x 10^-4^) and CSRP3 (OR 1.39, 95% CI 1.19 – 1.65, *p*= 1.29 x 10^-4^) (Figure 3C, eTable 8 in Supplement 1).

### Distinct protein modules associate with cognitive status in ALS

Next, we compared ALS-FTSD to non-FTSD-ALS. No significant differences in protein levels were found between the two groups (eFigure 6 in Supplement 1). However, when assessing how the identified protein modules differed between the phenotypes, we observed that the brown (r=0.23, p=0.004), immune-enriched turquoise (r=-0.19, p=0.02) and neurodevelopment-enriched yellow (r = -0.2, p = 0.009) modules were associated with ALS-FTSD (Figure 3B, eTable 7 in Supplement 1). Proteins included in the brown module were enriched for functions such as extracellular matrix organization (ECM) and cell adhesion (Figure 2, Supplement 2).

Increased levels of hub proteins within the ECM-enriched brown module, including NECTIN4 (OR 3.85, 95% CI 1.34–12.12, *p* = 0.034), JAM2 (OR 7.48, 95% CI 2.15–31.00, *p* = 0.032) and TGFBR2 (OR 4.28, 95% CI 1.32–15.68, *p* = 0.036), were associated with cognitive status in ALS (Figure 4A, eTable 9 in Supplement 1). A total of 21 hub proteins were identified, nine of them from the neurodevelopment-enriched yellow module and the hub proteins with the strongest associations to ALS-FTSD were TBCA (OR 0.4, 95% CI 0.21-0.72, *p=*0.032) and RWDD1(OR 0.31, 95% CI 0.13-0.72, *p=*0.032) (Figure 4A eTable 9 in Supplement 1).

**Figure 4.**
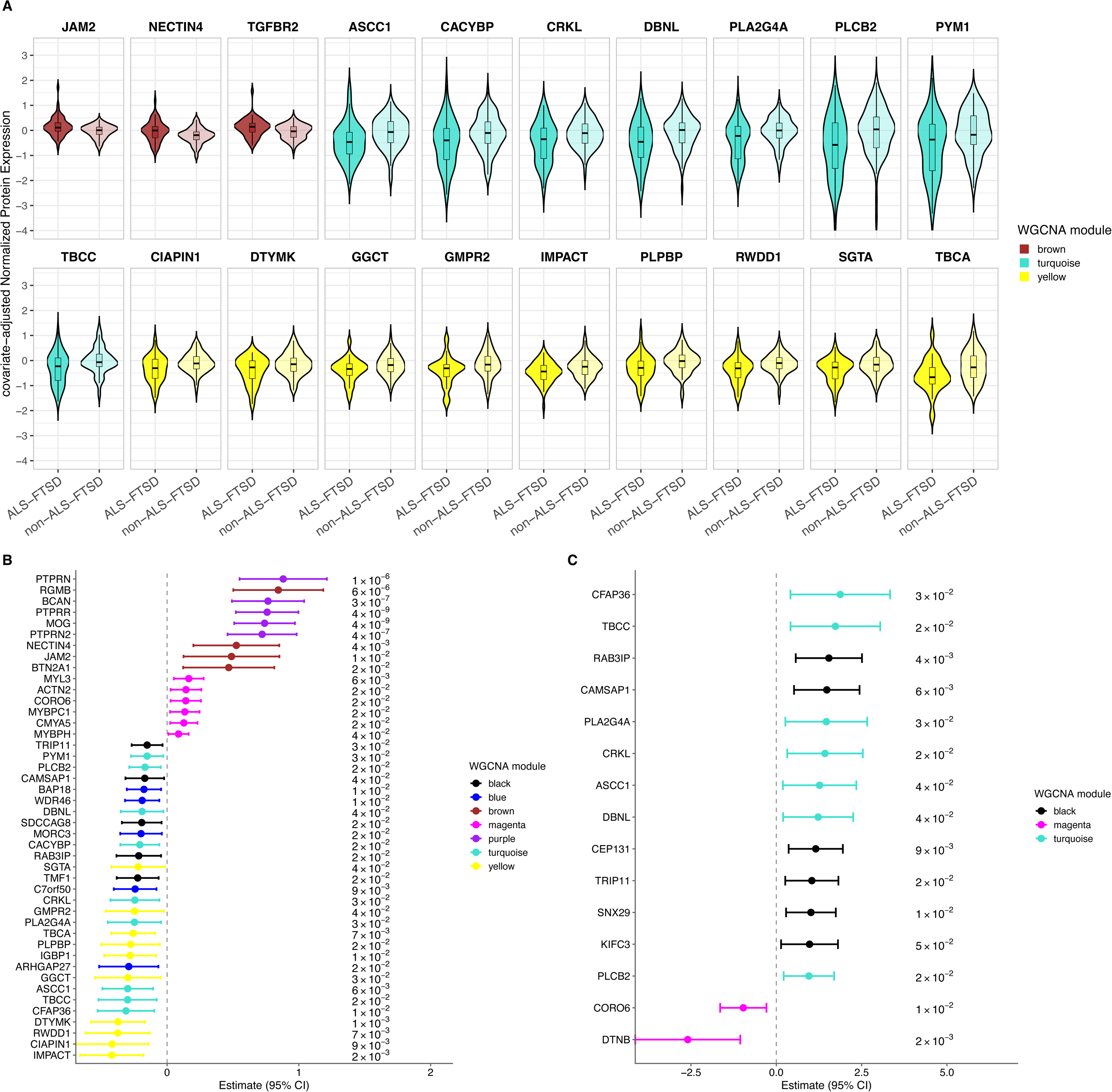
**A.** Violin plots of covariate-adjusted (median NPX, age and sex) normalized expression values of the 20 hub proteins significantly associated with ALS-FTSD (n=71) compared to non-ALS-FTSD (n=90), adjusted for ALSFRS-R score, BMI and site of symptom onset in logistic regression. Bar within each violin represents the median and interquartile range. Colors indicate WGCNA module membership. **B.** Forest plot with significant hub proteins for NEFL levels, determined with robust linear regression, adjusted for ALSFRS-R score, BMI and site of onset. FDR adjusted p-values shown to the right. **C.** ALSFRS-R score Hub Proteins. Beta estimates from robust linear regression models of 15 hub proteins significantly associated with ALSFRS-R score, adjusted for BMI and site of symptom onset. Protein abbrevations on y-axis, estimate on x-axis with 95%CI. Vertical dotted line marking 0. FDR-adjusted p-value on the right side of the plot. Coloring according to module assignment for each protein. Abbreviations: ALSFRS-R: amyotrophic lateral sclerosis functional rating scale; ALS-FTSD: amyotrophic lateral sclerosis-frontotemporal spectrum disorder; BMI: body mass index; CI: Confidence Interval; NPX: normalized protein expression; WGCNA: weighted gene correlation network analysis.

### Plasma NEFL levels are correlated with global alterations of plasma proteome in ALS

NEFL is a known marker of neuroaxonal injurywhich is increased in both plasma and CSF in ALS. Therefore, we assessed whether plasma NEFL levels correlated with global alterations of proteome defined by our protein modules.^15^ We observed that plasma NEFL was associated with six out of ten modules including muscle-enriched magenta, immune-enriched turquoise, and neurodevelopment-enriched yellow which we earlier identified as correlating with ALS diagnosis (Figure 3B, eTable 7 in Supplement 1). Notably, the synaptic signaling-enriched purple module had the strongest association to NEFL levels (r = 0.35, p = 5x10^-10^) (Figure 3B, eTable 7 in Supplement 1). We then examined whether individual proteins within the synaptic signaling-enriched purple module were associated with plasma NEFL levels. PTPRR (β = 0.75, 95 % CI 0.57–0.94, *p*= 9.92 x 10^-14^), MOG (β = 0.76, 95 % CI 0.56–0.96, *p*= 3.41 x 10^-12^) and PTPRN2 (β = 0.72, 95 % CI 0.5–0.94, *p*= 6.80 x 10^-10^) were associated with higher plasma NEFL levels (Figure 4B, eTable 10, eFigure 7 in Supplement 1). Interestingly, nine proteins within the muscle-enriched magenta module (CORO6, MYL3, MYBPC1, ACNT2, MYBPH, CMYA5, HSPB6. DTNB, CSRP3) that were increased in ALS patients when compared to controls, were also correlated with plasma NEFL levels (Figure 4B).

### ALSFRS-R score correlate with alterations in plasma proteome

We further sought to understand whether the detected protein modules can explain functional stages in ALS, by using ALSFRS-R scores. Module-trait association analysis revealed that protein abundances within the black (r=0.15, p=0.02) and turquoise (r=0.22, p=6 x 10^-4^) modules are subtly yet positively correlated with ALSFRS-R scores, indicating that increased levels of proteins included in these modules are related with higher ALSFRS-R score (Figure 3B, eTable 7 in Supplement 1). The muscle-enriched magenta module was negatively correlated with ALSFRS-R scores (r =−0.13, p=0.046), suggesting that increased levels of proteins in this module was correlated with lower ALSFRS-R scores (Figure 3B). A total of 15 hub proteins were significantly associated with ALSFRS-R score, after adjustment for onset site and BMI (Figure 4C, eTable 11 in Supplement 1). Modules associated with lower NEFL levels demonstrated the same direction of association as modules related to higher ALSFRS-R scores, indicating consistent expression patterns across traits and supporting the validity of the module construction (Figure 3B).

## Discussion

In this large plasma proteomic study of ALS, we applied differential abundance analysis, weighted gene correlation network analysis, and logistic regression modeling to identify proteins associated with disease and different clinical traits. Although 56 proteins were differentially abundant between ALS and controls, the network analysis revealed that the dominant signal originated from the muscle-enriched magenta module. This network, composed primarily of proteins involved in sarcomeric organization and contractile function, exhibited the strongest association with ALS diagnosis, and nearly three-quarters of all proteins increased in ALS belonged to this module (Figure 5). These network-level findings were supported by protein-level regression analyses, where muscle-enriched magenta hub proteins were consistently increased in ALS patients compared to controls.

**Figure 5.**
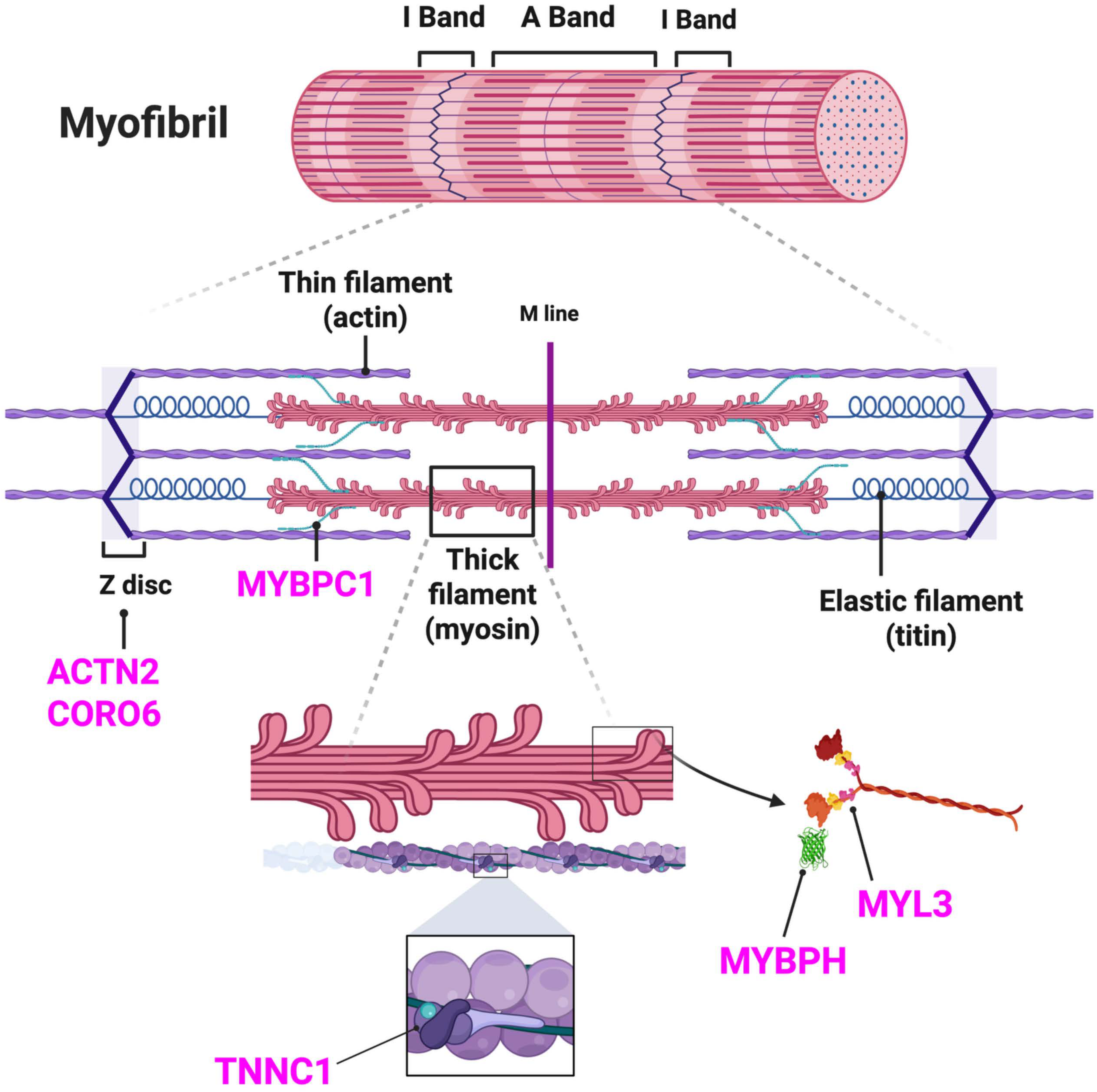
Myofibril illustration. ALS-specific hub proteins’ localization in myofibril and sarcomere structure. Magenta hub proteins are in magenta color. Illustration created with biorender.com.

Importantly, the biological relevance of the muscle-enriched magenta module extended beyond case–control discrimination, as it was also enriched in spinal-onset disease, where greater appendicular muscle mass is typically affected. Our findings are consistent with previous studies reporting increased muscle-related proteins in ALS, supporting the interpretation that peripheral muscle remodeling contributes substantially to the circulating proteomic profile.^21,37^ Our network approach adds granularity to previous observations by demonstrating that the alterations in proteome are not restrictive to single proteins, but to modules of highly correlated proteins that are associated with ALS diagnosis. This coherence strengthens biological interpretability and reinforces that muscle-linked proteins constitute a major component of the ALS plasma proteomic profile.

The onset site specific association further supports that the muscle-enriched magenta module captures the anatomical distribution of neuromuscular involvement, distinguishing it from generalized, non-specific systemic signals. Clinically, this distinction is meaningful, as spinal- and bulbar-onset ALS differ in progression and overall survival. At present, onset classification relies exclusively on clinical assessment, and a molecular correlate of the site of symptom onset has not been established. Our study suggests that the levels of muscle-derived proteins in plasma can discriminate between onset phenotypes, providing an objective biomarker for stratification in observational studies and clinical trials. If validated in independent cohorts, a composite “muscle involvement” score could complement clinical staging and functional measures, particularly in cases where phenotypic classification is ambiguous or lacking.

We observed that plasma NEFL levels were correlated with a considerable amount of the detected protein modules, most specifically the purple module which represents functions related to synaptic/neuronal signaling. This association supports the interpretation that increased NEFL reflects ongoing neuronal and synaptic injury. In addition, we observed a significant correlation, although weak, between plasma NEFL levels and the muscle-enriched magenta module, suggesting that neuroaxonal degeneration and peripheral muscle remodeling are biologically linked but partially dissociable processes.^11–13^

Separately, recent longitudinal work in pre-symptomatic pathogenic variant carriers identified rising NEFL prior to phenoconversion and showed that multi-protein panels can outperform NEFL alone for estimating time to phenoconversion.^23^ Together, these findings reinforce the role of NEFL as a marker of central neurodegeneration, while highlighting that it captures only part of the biological landscape of ALS, thereby supporting the added value of multi-protein panels that integrate complementary signals such as muscle remodeling. This motivates a framework of complementary biomarkers, where NEFL and muscle-associated markers jointly characterize disease state and heterogeneity rather than serving as redundant measures.

While the muscle-enriched magenta module dominated ALS vs control and onset-site associations, cognitive status was instead mapped to the brown module, enriched for extracellular matrix organization and cell adhesion. This suggests that cognitive phenotypes in ALS might have different biological underpinnings than site of onset which were predominantly related to muscle remodeling.

The differential mapping of plasma proteome signatures across the clinical traits supports a model in which ALS heterogeneity is multi-dimensional. One axis reflects neuromuscular remodeling and altered levels of muscle proteins, whereas another aligns more closely to processes implicated in cognitive/FTD-spectrum features. This consolidate the view that ALS is a biologically heterogeneous disorder and its sub-phenotypes may be better represented by pathway-level signatures than by single biomarkers.^1,2^

Two recent studies illustrate the field’s momentum toward blood-based proteomic biomarkers.^22,23^ Chia et al, using Olink Explore 3072, identified 33 differentially abundant proteins and built a machine-learning model with high diagnostic accuracy, and interpreted pathway-level signals implicating skeletal muscle, nerves, and energy metabolism.^22^ Notably, several ALS associated proteins identified in our data, including CMYA5 and TNNC1 were not reported in the study by Chia et al, but were identified by Ran et al, using Olink Explore HT, a finding we replicate here.^22,23^

Our correlation network analysis complements these advances by showing that muscle-related changes are captured in a module (magenta) strongly associated with ALS, while cognition maps to a distinct ECM-enriched module (brown), with TGFBR2 and NECTIN4 as prominent features. Our approach offers two advantages in understanding the disease biology. First, it offers interpretability where correlating proteins reflect coordinated biological processes than isolated univariate tests. Second, the identified protein modules allow to correlate distinct clinical traits with biological functions represented by each of these modules enabling biology-informed stratification beyond the case-control contrasts.

The coherence between our module-level signals and patterns reported in independent Olink, SOMAscan, and mass-spectrometry studies suggests that certain pathway signatures, particularly muscle-related and adhesion processes, are reproducibly observed across proteomic platforms in ALS. This cross-technology consistency strengthens the biological validity of these signals and supports their relevance as candidate biomarkers.^22,23,38^

## Limitations

Key limitations should be acknowledged. First, the study is cross-sectional for most trait associations, limiting causal inference and the ability to determine whether module differences precede clinical transitions. Second, plasma proteomics could be influenced by peripheral tissues; muscle-related signals may dominate and could partially reflect disease stage or systemic factors not fully captured by covariates. Third, external replication of module–trait associations are needed, particularly for within-ALS analyses (spinal vs bulbar; ALS-FTSD vs non-ALS-FTSD). While we adjusted for covariates, some systemic factors such as inflammation, physical activity, cardiovascular strain and co-morbidities were not included in this study. Future studies with longitudinal proteomics data accompanied with more clinical data, such as co-morbidities, socioeconomic status and lifestyle, would give further insight to the general biological processes and pathogenic drivers of this heterogenous disease.

## Conclusion

In summary, network-based plasma proteomics revealed a dominant muscle/sarcomere module strongly associated with ALS and sensitive to onset phenotype, alongside distinct modules linked to cognition and neuroaxonal injury. Together these findings support a multi-axis model of ALS heterogeneity in which neuromuscular remodeling and neuroaxonal injury represent related but partially separable biological dimensions, with clear implications for patient stratification and biomarker panel design.

Our results reinforce the emerging view that ALS is not driven by a single dominant pathway, but instead reflects multiple partly independent processes, including neuroaxonal injury, muscule remodeling, and extracellular matrix/vascular signaling, each of which may contribute differently across patients.

## Data Availability

All data produced in the present study are available upon reasonable request to the authors

## Acknowledgements

We would like to thank all the participants for their invaluable contribution to this work. Further we thank all the team members working with collecting samples and data. Special thanks to Jenny Hellqvist for her precious help with data collection. This study was funded by Region Stockholm, ALS-fonden and Börje Salming ALS foundation.

## Ethics approval and consent to participate

The study was conducted in accordance with the Declaration of Helsinki. An informed consent to participate in research was obtained from each participant. The study was reviewed and approved by the Swedish Ethical Review Authority (diary numbers 2014/1815-31/4, 2017/1895-31, 2017/828-32, 2018/1605-31, 2018/2409, 2019-02338, 2021-06397-02, 2024-07786-02).

## Availability of data and materials

The clinical dataset used and analyzed during the current study are available from the corresponding author upon reasonable request and establishment of data transfer agreement.

## Author Contributions

Concept and design: Månberg, Ingre

Acquisition, analysis, or interpretation of data: Azizi, Aksoylu, Foucher, Álvez, Öijerstedt, Edfors, Bergström, Månberg, Ingre, Juto, Press, Samuelsson, Kläppe

Drafting of the manuscript: Azizi, Aksoylu, Álvez, Öijerstedt, Månberg, Ingre

Critical review of the manuscript for important intellectual content: Azizi, Aksoylu, Álvez, Foucher, Juto, Seitz, Press, Samuelsson, Kläppe, Uhlén, Edfors, Bergström, Fang, Nilsson, Öijerstedt, Månberg, Ingre

Statistical analysis: Azizi, Aksoylu, Álvez, Öijerstedt

Obtained funding: Ingre, Månberg

Administrative, technical, or material support: Azizi, Aksoylu, Álvez, Öijerstedt, Bergström Månberg, Ingre, Seitz

Supervision: Ingre, Månberg

## Abbreviations table

ACTN2: Alpha-actinin-2
ALS: Amyotrophic lateral sclerosis
ALS-FTSD: Amyotrophic lateral sclerosis-frontotemporal spectrum disorder
ALSFRS-R: Amyotrophic Lateral Sclerosis Functional Rating Scale-Revised
ANT3: Antithrombin III
ASCC1: Activating signal cointegrator 1 complex subunit 1
AUC: Area under the receiver operating characteristic curve
BMI: Body mass index
CACYBP: Calcyclin-binding protein
CFAP36: Cilia and flagella associated protein 36
CIAPIN1: Cytokine induced apoptosis inhibitor 1
CMYA5: Cardiomyopathy associated 5
CORO6: Coronin 6
CRKL: CRK-like proto-oncogene, adaptor protein
CSF: Cerebrospinal fluid
CSRP3: Cysteine and glycine-rich protein 3
DBNL: Drebrin-like
DTNB: Dystrobrevin beta
DTYMK: Deoxythymidylate kinase
ECAS: Edinburgh Cognitive and Behavioral ALS Screen
EMG: Electromyography
FDR: False discovery rate
FTD: Frontotemporal dementia
GGCT: Gamma-glutamylcyclotransferase
GMPR2: Guanosine monophosphate reductase 2
GO: Gene Ontology
GRAP2: GRB2-related adaptor protein 2
HSPB6: Heat shock protein family B (small) member 6
IGBP1: Immunoglobulin binding protein 1
IMPACT: IMPACT RWD domain protein
IQR: Interquartile range
MM: Module membership
MYBPC1: Myosin-binding protein C1
MYBPH: Myosin-binding protein H
MYL3: Myosin light chain 3
NEFL: Neurofilament light chain
NPX: Normalized Protein eXpression
PEA: Proximity Extension Assay
PLA2G4A: Phospholipase A2 group IVA
PLCB2: Phospholipase C beta 2
PLPBP: Pyridoxal phosphate binding protein
PYM1: PYM1 homolog, exon-junction complex recycling factor
QC: Quality control
RWDD1: RWD domain containing 1
SGTA: Small glutamine rich tetratricopeptide repeat containing alpha
TBCA: Tubulin folding cofactor A
TBCC: Tubulin folding cofactor C
TNNC1: Troponin C1, slow skeletal and cardiac type
WGCNA: Weighted gene correlation network analysis

