## Supplement 1 for "Muscle proteins in plasma associate to distinguished phenotypes in amyotrophic lateral sclerosis"

### Supplemental Online Content

**eFigure 1.** PCA before covariate correction

**eFigure 2.** Scale−free topology analysis

**eFigure 3.** Mean Connectivity Analysis

**eFigure 4.** Protein dendrogram and module colors

**eFigure 5.** Clustering dendrogram of module eigengenes

**eFigure 6.** Volcano plot ALS−FTSD versus non−ALS−FTSD

**eFigure 7.** Correlation between neurofilament light chain and selected correlation proteins in ALS patients

**eTable 1.** Differentially expressed proteins ALS versus control

**eTable 2.** Module colors ALS versus control

**eTable 3.** Hub proteins per correlation module

**eTable 4.** Module-trait association ALS versus control from weighted gene correlation network analysis

**eTable 5.** Logistic regression ALS versus controls

**eTable 6.** Differentially expressed proteins Spinal versus Bulbar onset

**eTable 7.** Module-trait association ALS-specific traits from weighted gene correlation network analysis

**eTable 8.** Logistic regression Spinal versus Bulbar onset

**eTable 9.** Logistic regression ALS-FTSD versus non-ALS-FTSD

**eTable 10.** Robust linear regression NEFL

**eTable 11.** Robust linear regression ALSFRS-R score

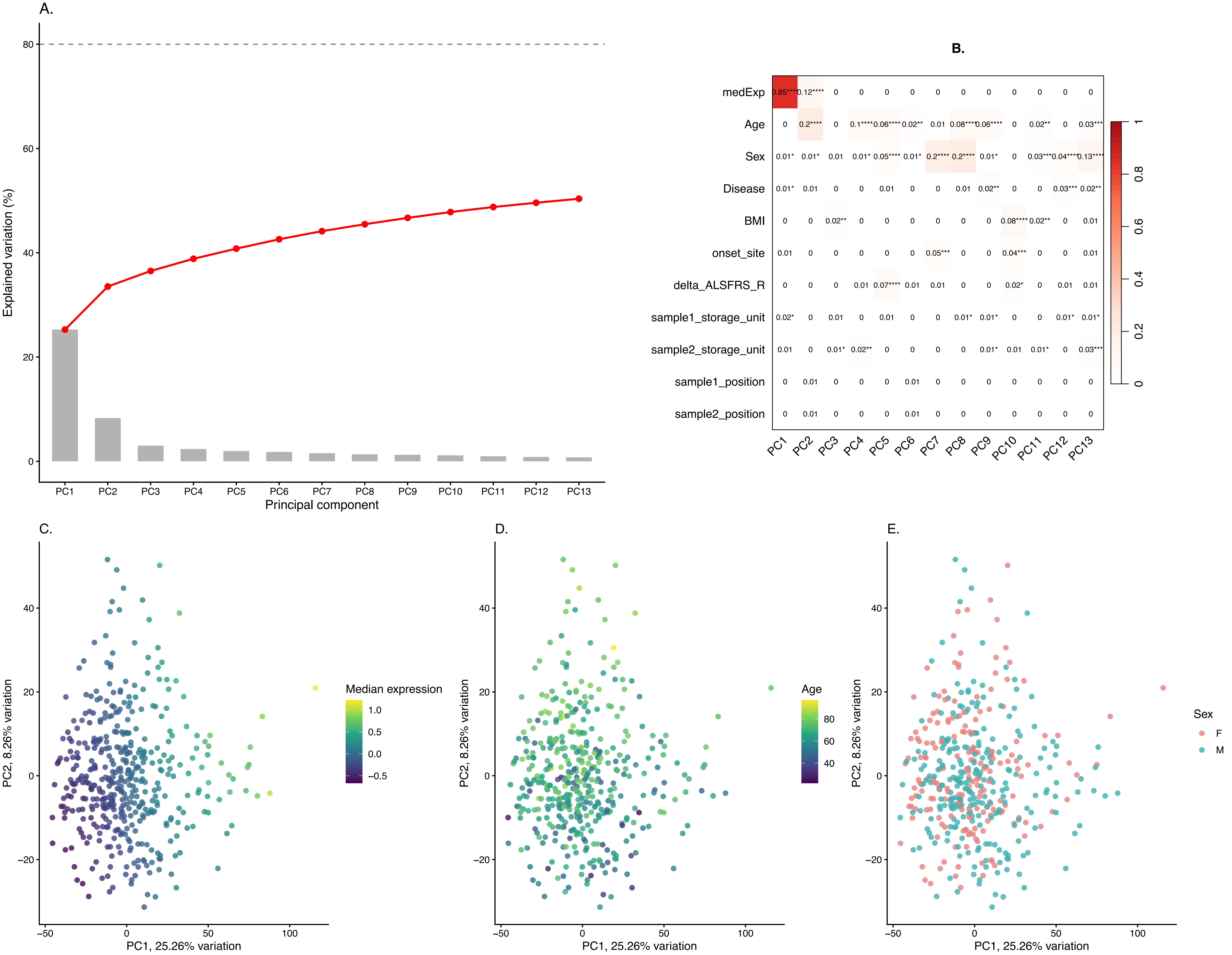


**eFigure 1. Principal component analysis of plasma proteome. A.** Scree plot showing the variance explained (y–axis) by each principal component (PC) (x–axis). Cumulative sum of variance is represented by the red line, explained by all PCs. Cumulative variance threshold (80%) is represented by dashed horizontal line whereas the vertical line represents the PCs that explain highest cumulative amount of variation in the data. **B.** Heatmap displaying Pearson correlations between principal component eigenvalues (columns) and technical and clinical covariates (rows). Cell color reflects the magnitude of the absolute correlation, and coefficients are shown numerically within each cell. Significance is indicated by asterisks (none: p > 0.05; *: p < 0.05; **: p < 0.01; ***: p < 0.0001; ****: p ~ 0). **C, D and E.** Biplots of PC1 (x–axis; 25.26% of variance explained) and PC2 (y–axis; 8.26% of variance explained), with samples colored by median NPX expression, age, and sex, respectively.

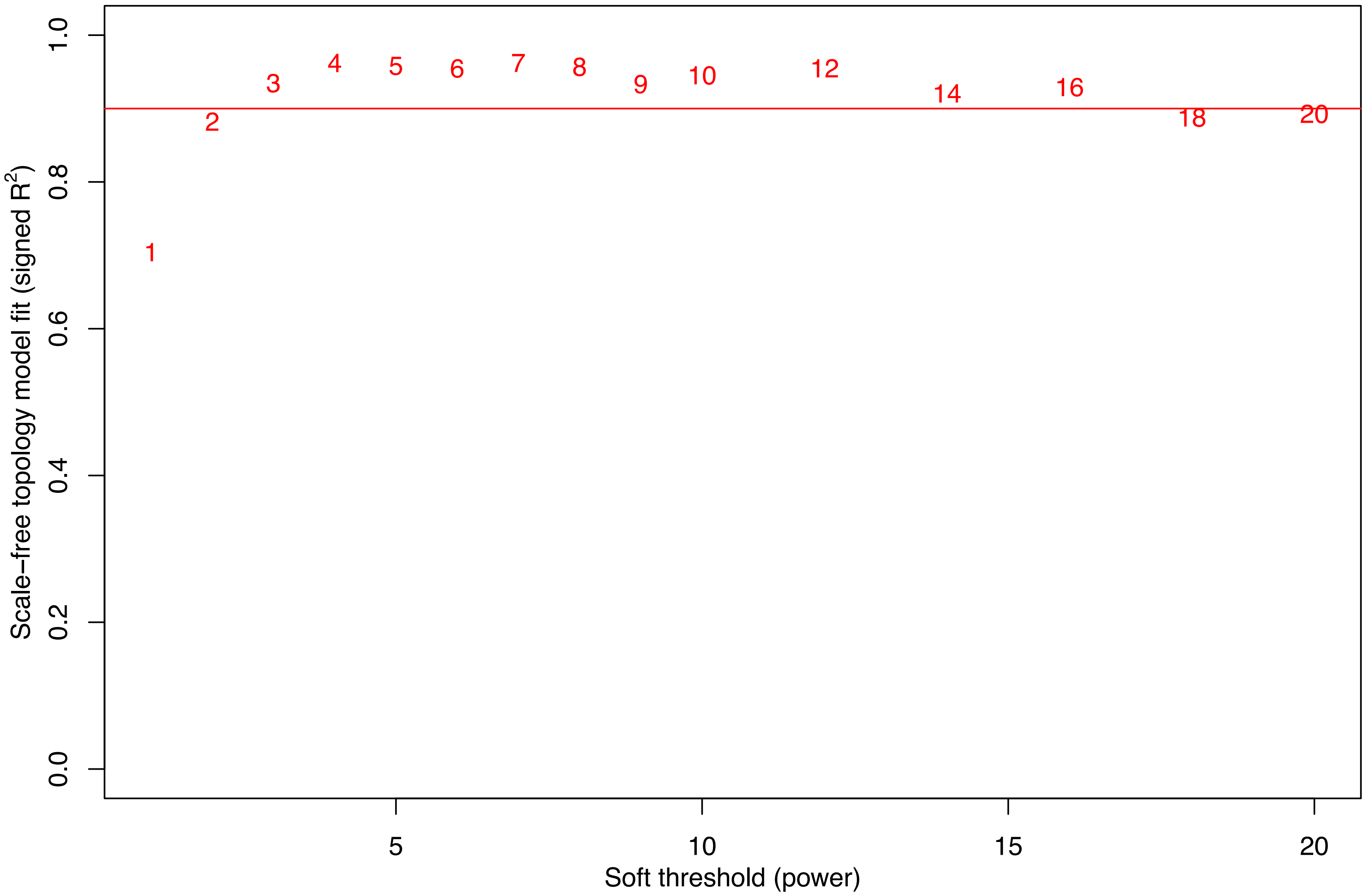


**eFigure 2. Scale−free topology analysis.** Evaluation of the scale−free fit index (y−axis) across soft−thresholding powers. The red line indicates the $R^2$ cut−off of 0.9, used to identify the optimal power for network construction to ensure a scale−free topology.

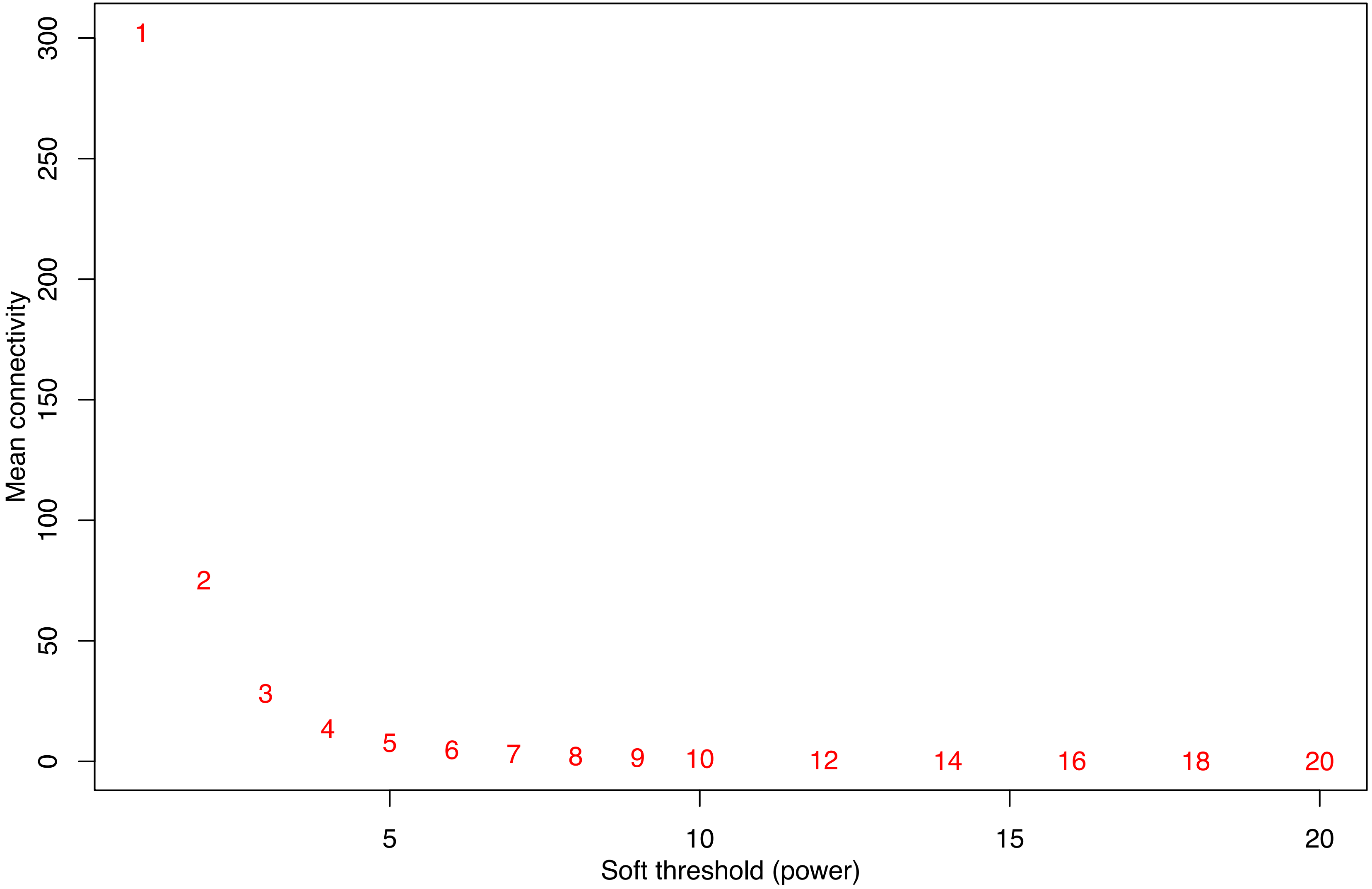


**eFigure 3. Mean Connectivity Analysis.** Analysis of the mean connectivity (y−axis) across various soft−thresholding powers (x−axis) for the protein network. Numbers in red indicate the specific power evaluated to identify the optimal scale−free topology.

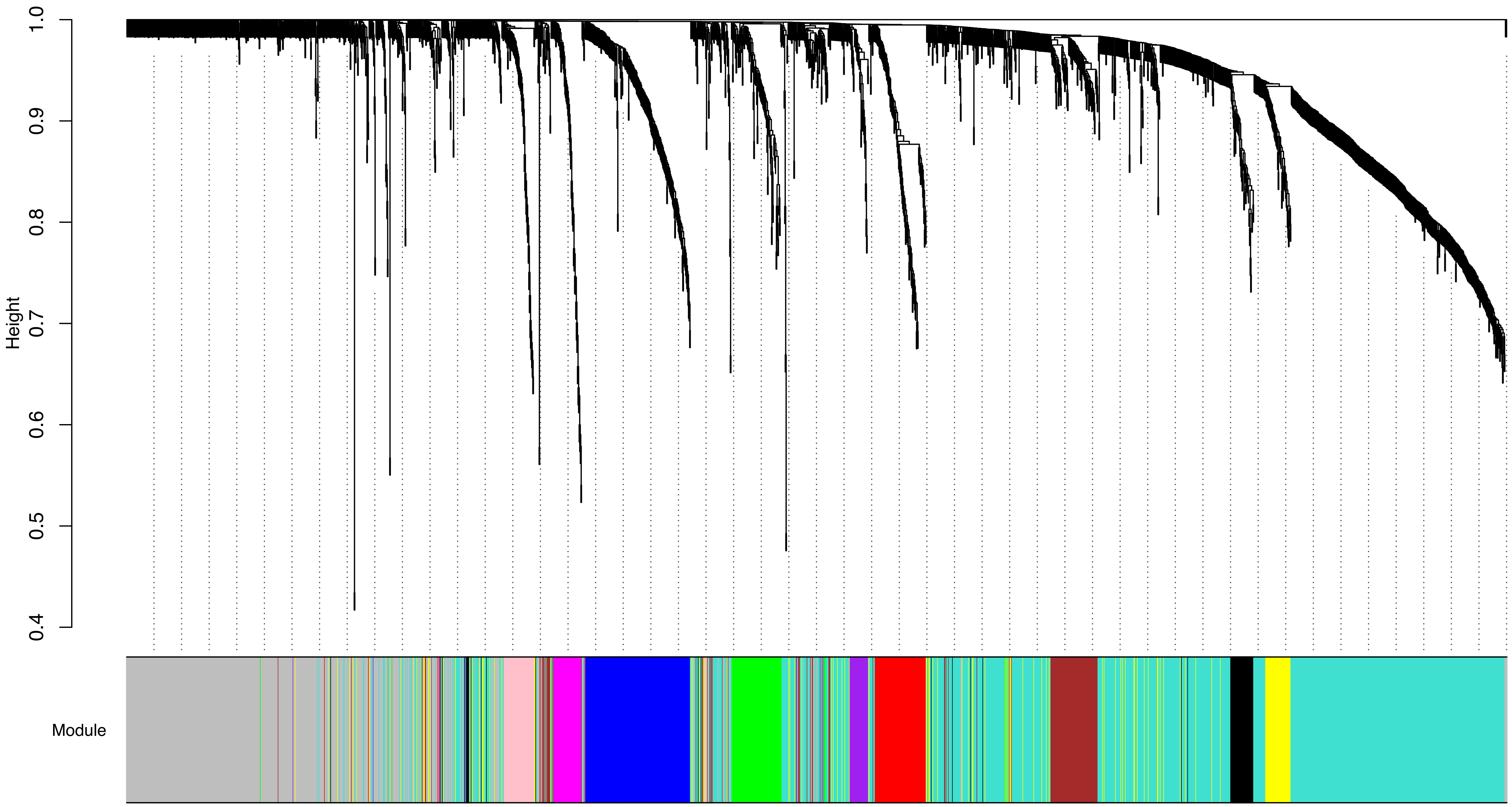


**eFigure 4. Protein dendrogram and module colors**. Hierarchical clustering of proteins into 11 modules based on a topological overlap matrix. Each color row represents a distinct module,while the 'grey' module contains unassigned proteins. Dendrogram branch and guide hanging (0.03 and 0.05, respectively) were adjusted for visual clarity. Modules created: black, blue, brown, green, magenta, pink, purple, red, turquoise and yellow

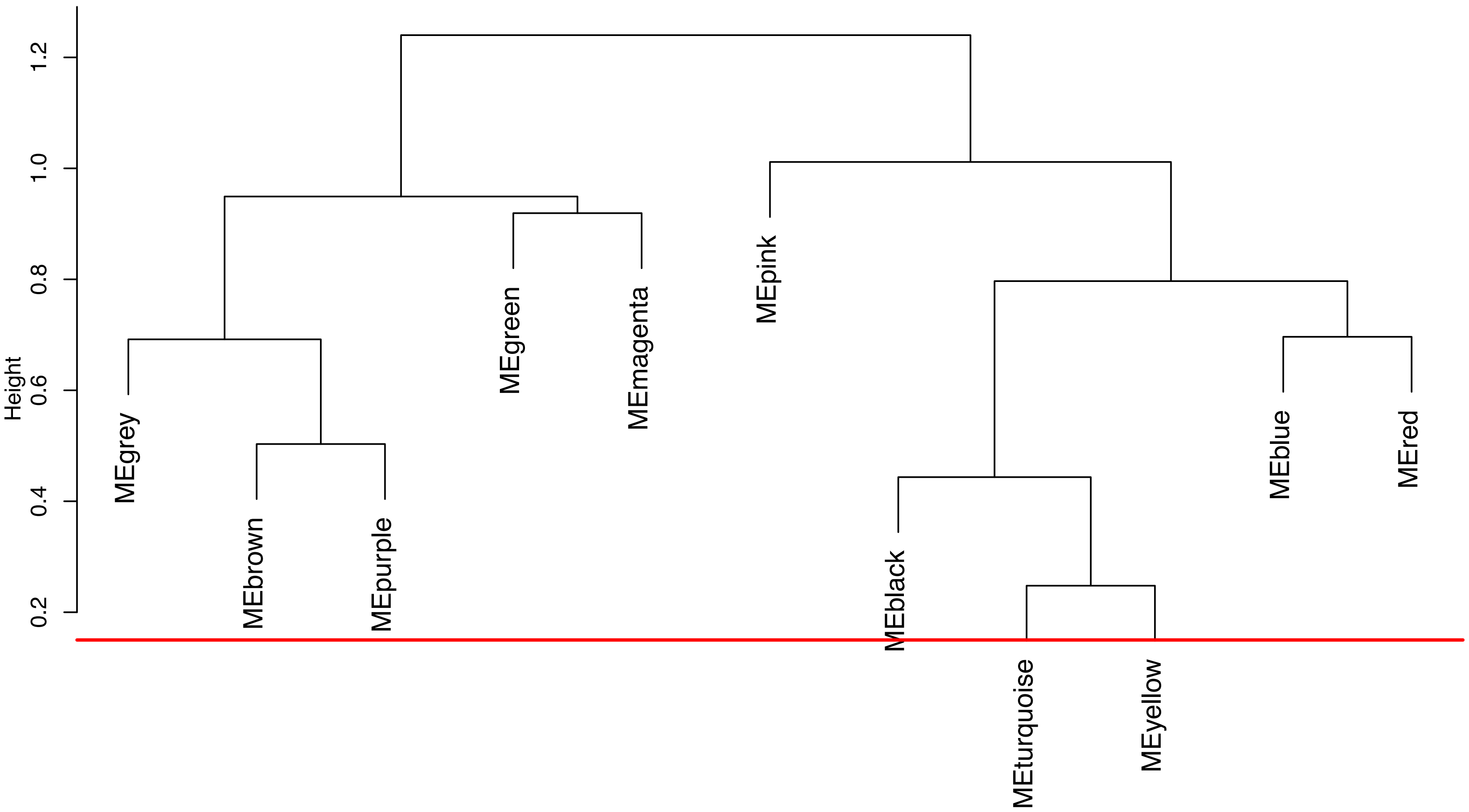


**eFigure 5. Clustering dendrogram of module eigengenes.** Hierarchical clustering of module eigengenes based on dissimilarity (1 – correlation). The red line at height 0.15 indicates the threshold used for merging highly correlated modules to ensure distinct correlation patterns. None were eligble for merging.

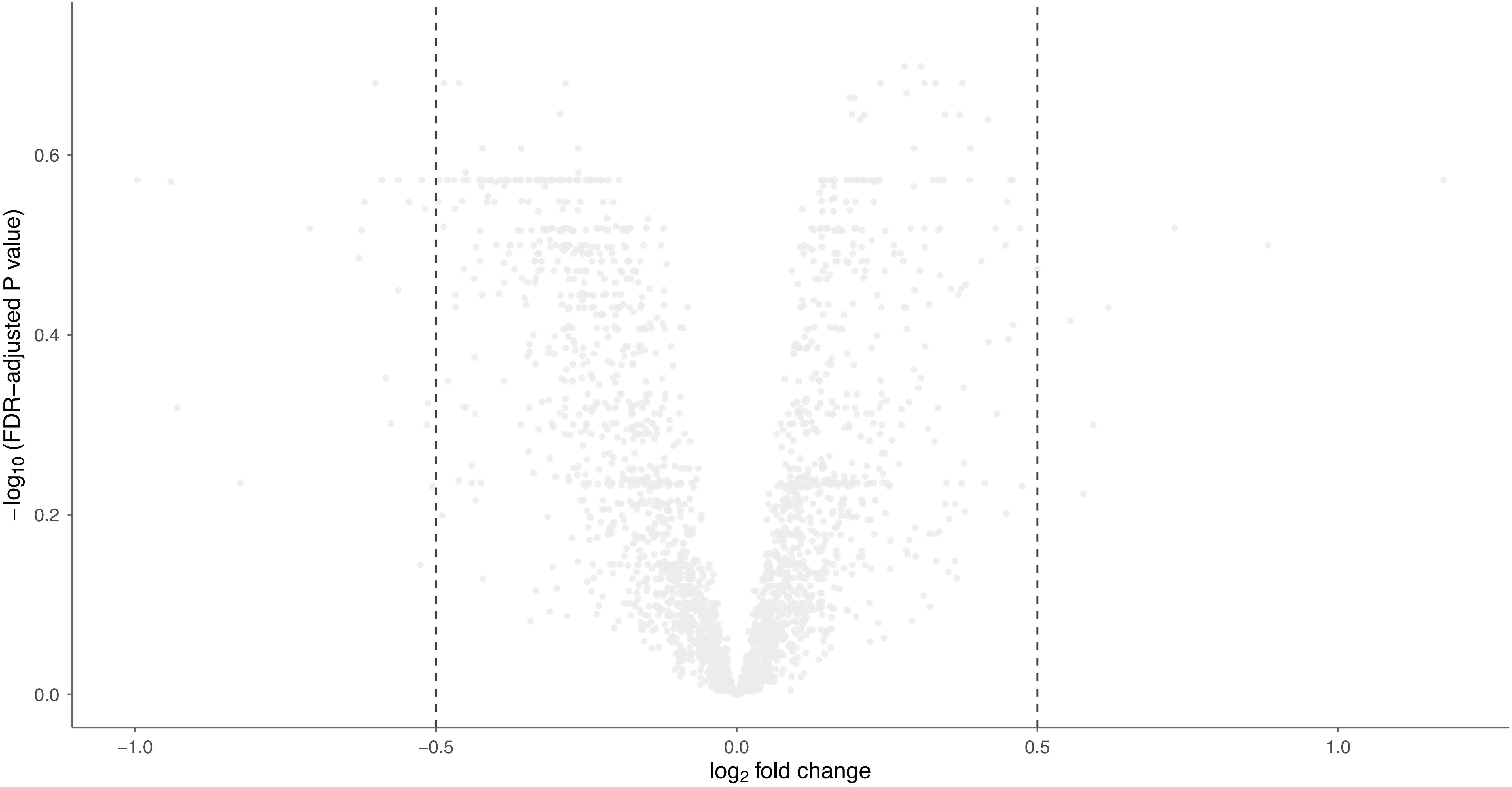


**eFigure 6. ALS−FTSD versus non−ALS−FTSD** Protein level comparison between ALS−FTSD (n=71) and non−ALS−FTSD (n=90), adjusted for ALSFRS−R score, BMI and site of symptom onset. Dashed lines indicate inclusion thresholds (horizontal: FDR adjusted p−value< 0.05; vertical: $|\log_2$−fold change$| > 0.5$), marking the inclusion criteria. No protein met inclusion criteria.
Abbreviations: ALSFRS−R: amyotrophic lateral sclerosis functional rating scale; ALS−FTSD: amyotrophic lateral sclerosis−frontotemporal spectrum disorder; BMI: body mass index

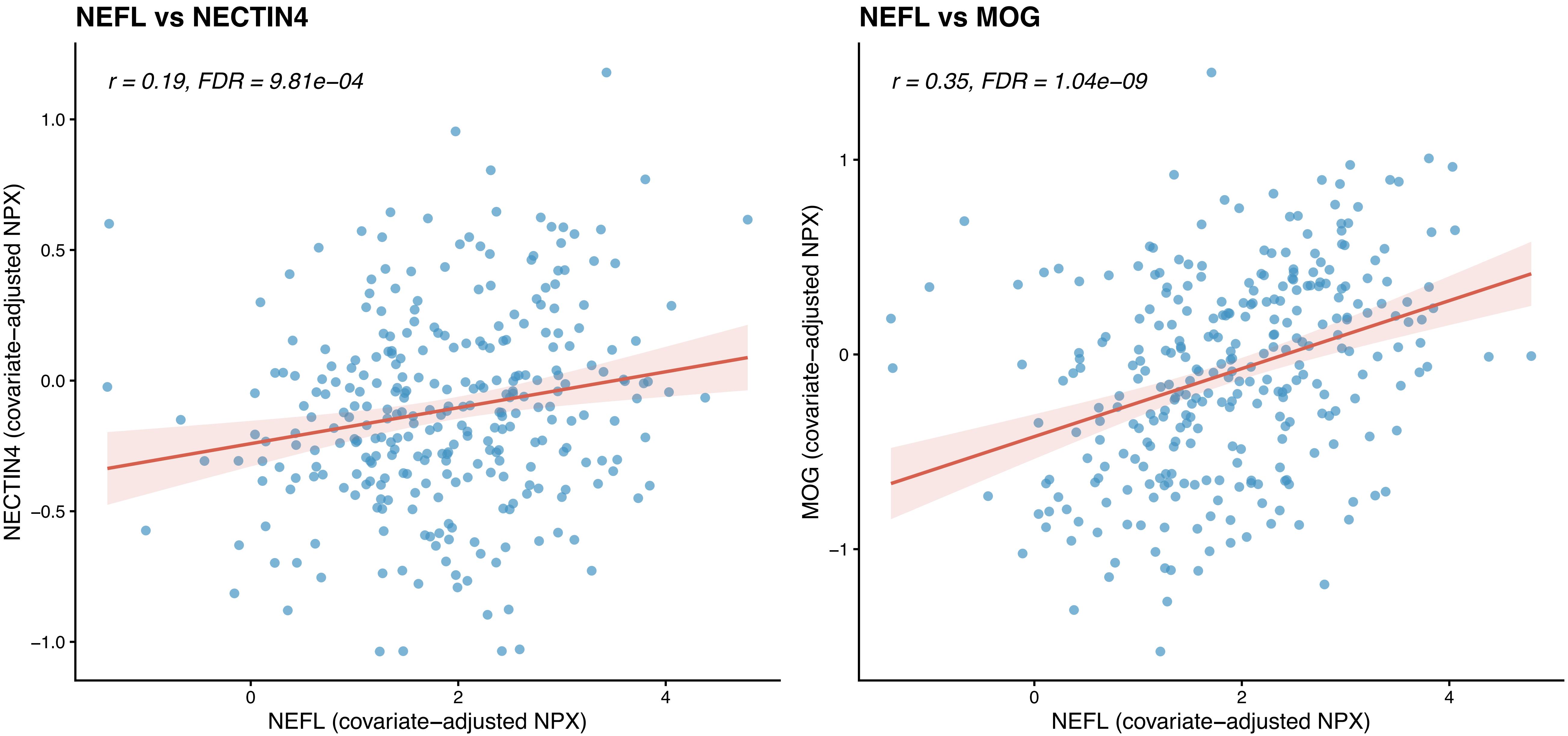


**eFigure 7. Correlation between neurofilament light chain and selected co−expressed proteins in ALS patients.** Scatter plots show Pearson correlations between NEFL and NECTIN4 (left, brown module) and NEFL and MOG (right, purple module) in ALS patients. Each point represents one individual. Values on both axes are covariate−adjusted NPX (Normalized Protein eXpression). The red line indicates the linear regression fit with 95% confidence interval (shaded). Correlation coefficients and FDR−adjusted P values are shown in each panel.

**eTable 1. Differentially expressed proteins ALS versus control.** All proteins meeting FDR < 0.05 are displayed. Proteins meeting criteria FDR <0.05 and |log2 fold change| > 0.5 are in bold. P values are FDR-adjusted (Benjamini–Hochberg). Ordered with ascending FDR-adjusted p-value.

| Protein | UniProt ID | log2 fold change | FDR-adjusted P value | WGCNA module |
|---|---|---|---|---|
| **NEFL** | **P07196** | **2.34** | **$5.23 \times 10^{-37}$** | **-** |
| **MEGF10** | **Q96KG7** | **0.97** | **$2.82 \times 10^{-18}$** | **magenta** |
| **HS6ST2** | **Q96MM7** | **0.92** | **$7.38 \times 10^{-18}$** | **magenta** |
| **CSRP3** | **P50461** | **2.29** | **$2.54 \times 10^{-14}$** | **magenta** |
| **MYBPC1** | **Q00872** | **1.63** | **$4.11 \times 10^{-14}$** | **magenta** |
| **ACTN2** | **P35609** | **1.46** | **$7.40 \times 10^{-13}$** | **magenta** |
| **MYBPH** | **Q13203** | **1.96** | **$1.75 \times 10^{-12}$** | **magenta** |
| **CORO6** | **Q6QEF8** | **1.39** | **$2.28 \times 10^{-12}$** | **magenta** |
| **EDA2R** | **Q9HAV5** | **0.91** | **$3.08 \times 10^{-11}$** | **magenta** |
| **MYBPC2** | **Q14324** | **1.58** | **$3.45 \times 10^{-11}$** | **magenta** |
| **HSPB6** | **O14558** | **0.66** | **$5.32 \times 10^{-11}$** | **magenta** |
| **TNNC1** | **P63316** | **1.78** | **$9.47 \times 10^{-11}$** | **magenta** |
| **DTNB** | **O60941** | **0.62** | **$1.88 \times 10^{-10}$** | **magenta** |
| **CMYA5** | **Q8N3K9** | **1.35** | **$2.36 \times 10^{-10}$** | **magenta** |
| **RBFOX3** | **A6NFN3** | **0.78** | **$4.12 \times 10^{-10}$** | **magenta** |
| **AFAP1L1** | **Q8TED9** | **0.55** | **$1.28 \times 10^{-9}$** | **magenta** |
| **ART3** | **Q13508** | **-0.54** | **$9.98 \times 10^{-9}$** | **brown** |
| **HPCAL4** | **Q9UM19** | **0.74** | **$1.16 \times 10^{-8}$** | **magenta** |
| **MYOM3** | **Q5VTT5** | **1.26** | **$1.16 \times 10^{-8}$** | **magenta** |
| **MYL3** | **P08590** | **1.08** | **$4.93 \times 10^{-8}$** | **magenta** |
| COMP | P49747 | -0.44 | $5.43 \times 10^{-8}$ | grey |
| **SYNM** | **O15061** | **0.73** | **$1.44 \times 10^{-7}$** | **magenta** |
| TNFRSF12A | Q9NP84 | 0.45 | $3.02 \times 10^{-7}$ | magenta |
| **DUSP29** | **Q68J44** | **1.10** | **$5.64 \times 10^{-7}$** | **magenta** |
| **DMD** | **P11532** | **0.52** | **$1.08 \times 10^{-6}$** | **magenta** |
| **NEB** | **P20929** | **0.91** | **$1.08 \times 10^{-6}$** | **magenta** |
| **GH1** | **P01241** | **1.79** | **$1.27 \times 10^{-6}$** | **grey** |
| **NOS1** | **P29475** | **0.61** | **$3.93 \times 10^{-6}$** | **magenta** |

| Protein | UniProt ID | log2 fold change | FDR-adjusted P value | WGCNA module |
|---|---|---|---|---|
| **MYH1** | **P12882** | **1.35** | **$6.46 \times 10^{-6}$** | **magenta** |
| **ALDH3A1** | **P30838** | **1.54** | **$8.87 \times 10^{-6}$** | **turquoise** |
| ITGB6 | P18564 | -0.37 | $9.81 \times 10^{-6}$ | grey |
| **FABP3** | **P05413** | **0.52** | **$1.01 \times 10^{-5}$** | **magenta** |
| IGFBP6 | P24592 | -0.31 | $1.01 \times 10^{-5}$ | brown |
| **CALCA** | **P01258** | **0.74** | **$1.18 \times 10^{-5}$** | **magenta** |
| CHCHD10 | Q8WYQ3 | 0.40 | $6.63 \times 10^{-5}$ | magenta |
| CD276 | Q5ZPR3 | 0.41 | $1.02 \times 10^{-4}$ | turquoise |
| **SORBS1** | **Q9BX66** | **0.51** | **$1.22 \times 10^{-4}$** | **magenta** |
| LMNB2 | Q03252 | 0.32 | $1.67 \times 10^{-4}$ | turquoise |
| PDCD1 | Q15116 | 0.45 | $2.58 \times 10^{-4}$ | turquoise |
| TNFRSF8 | P28908 | 0.40 | $2.58 \times 10^{-4}$ | turquoise |
| **PRPH** | **P41219** | **0.51** | **$3.21 \times 10^{-4}$** | **grey** |
| USP28 | Q96RU2 | 0.44 | $4.84 \times 10^{-4}$ | magenta |
| HS1BP3 | Q53T59 | -0.41 | $5.48 \times 10^{-4}$ | turquoise |
| SUSD4 | Q5VX71 | 0.35 | $6.12 \times 10^{-4}$ | turquoise |
| NUMB | P49757 | -0.47 | $9.24 \times 10^{-4}$ | turquoise |
| **KLK4** | **Q9Y5K2** | **0.60** | **$9.67 \times 10^{-4}$** | **turquoise** |
| ITGB5 | P18084 | -0.26 | $1.19 \times 10^{-3}$ | grey |
| NRCAM | Q92823 | 0.25 | $1.19 \times 10^{-3}$ | magenta |
| ADAMTS1 | Q9UHI8 | 0.23 | $1.20 \times 10^{-3}$ | turquoise |
| **OPHN1** | **O60890** | **-0.66** | **$1.64 \times 10^{-3}$** | **turquoise** |
| NT5C1A | Q9BXI3 | 0.27 | $2.04 \times 10^{-3}$ | turquoise |
| EIF2B4 | Q9UI10 | -0.42 | $2.32 \times 10^{-3}$ | yellow |
| **GCG** | **P01275** | **-0.81** | **$2.32 \times 10^{-3}$** | **grey** |
| OGA | O60502 | -0.37 | $2.40 \times 10^{-3}$ | turquoise |
| RGMA | Q96B86 | -0.27 | $2.40 \times 10^{-3}$ | brown |
| **CCK** | **P06307** | **-0.75** | **$2.48 \times 10^{-3}$** | **grey** |
| **SSC4D** | **Q8WTU2** | **-0.85** | **$3.03 \times 10^{-3}$** | **grey** |
| ARHGEF1 | Q92888 | -0.44 | $3.23 \times 10^{-3}$ | turquoise |
| TSC22D3 | Q99576 | -0.46 | $3.28 \times 10^{-3}$ | turquoise |

| Protein | UniProt ID | log2 fold change | FDR-adjusted P value | WGCNA module |
|---|---|---|---|---|
| **UFD1** | **Q92890** | **-0.72** | **$3.39 \times 10^{-3}$** | **turquoise** |
| NCR3LG1 | Q68D85 | 0.25 | $4.48 \times 10^{-3}$ | turquoise |
| CXCL10 | P02778 | 0.47 | $4.68 \times 10^{-3}$ | yellow |
| **KRT5** | **P13647** | **-0.57** | **$4.68 \times 10^{-3}$** | **grey** |
| VASP | P50552 | -0.44 | $4.68 \times 10^{-3}$ | turquoise |
| EIF4B | P23588 | -0.39 | $5.49 \times 10^{-3}$ | yellow |
| PEPD | P12955 | -0.21 | $5.49 \times 10^{-3}$ | grey |
| TPPP3 | Q9BW30 | -0.47 | $5.71 \times 10^{-3}$ | grey |
| AXIN1 | O15169 | -0.39 | $5.77 \times 10^{-3}$ | turquoise |
| MANSC1 | Q9H8J5 | 0.22 | $5.77 \times 10^{-3}$ | brown |
| RTN4IP1 | Q8WWV3 | -0.43 | $5.77 \times 10^{-3}$ | turquoise |
| BCR | P11274 | -0.38 | $6.15 \times 10^{-3}$ | turquoise |
| AKAP2 | Q9Y2D5 | 0.21 | $6.24 \times 10^{-3}$ | turquoise |
| AMIGO2 | Q86SJ2 | 0.10 | $6.74 \times 10^{-3}$ | turquoise |
| LCAT | P04180 | -0.14 | $7.12 \times 10^{-3}$ | grey |
| KLK8 | O60259 | -0.25 | $7.30 \times 10^{-3}$ | brown |
| ARHGEF12 | Q9NZN5 | -0.42 | $7.41 \times 10^{-3}$ | turquoise |
| BANK1 | Q8NDB2 | -0.46 | $7.57 \times 10^{-3}$ | turquoise |
| IKBKG | Q9Y6K9 | -0.30 | $7.57 \times 10^{-3}$ | turquoise |
| CCL22 | O00626 | 0.31 | $8.62 \times 10^{-3}$ | turquoise |
| USP8 | P40818 | -0.44 | $8.78 \times 10^{-3}$ | turquoise |
| CD80 | P33681 | 0.24 | $9.06 \times 10^{-3}$ | turquoise |
| ASPSCR1 | Q9BZE9 | -0.26 | $9.15 \times 10^{-3}$ | yellow |
| SNX2 | O60749 | -0.41 | $9.15 \times 10^{-3}$ | turquoise |
| DEFA1 | P59665 | 0.38 | $9.75 \times 10^{-3}$ | turquoise |
| CAPN3 | P20807 | 0.49 | $9.78 \times 10^{-3}$ | magenta |
| EML4 | Q9HC35 | -0.40 | $9.78 \times 10^{-3}$ | turquoise |
| IPCEF1 | Q8WWN9 | -0.40 | $1.01 \times 10^{-2}$ | turquoise |
| CHUK | O15111 | -0.42 | $1.03 \times 10^{-2}$ | yellow |
| NOL3 | O60936 | 0.27 | $1.11 \times 10^{-2}$ | magenta |
| S100A12 | P80511 | 0.39 | $1.11 \times 10^{-2}$ | blue |

| Protein | UniProt ID | log2 fold change | FDR-adjusted P value | WGCNA module |
|---|---|---|---|---|
| ST13 | P50502 | -0.28 | $1.11 \times 10^{-2}$ | red |
| IMPACT | Q9P2X3 | -0.27 | $1.21 \times 10^{-2}$ | yellow |
| ITIH3 | Q06033 | 0.25 | $1.21 \times 10^{-2}$ | turquoise |
| DDR1 | Q08345 | 0.15 | $1.22 \times 10^{-2}$ | turquoise |
| PADI2 | Q9Y2J8 | 0.62 | $1.28 \times 10^{-2}$ | magenta |
| SCARF2 | Q96GP6 | 0.21 | $1.28 \times 10^{-2}$ | brown |
| IAH1 | Q2TAA2 | -0.25 | $1.31 \times 10^{-2}$ | yellow |
| **CCL26** | **Q9Y258** | **0.52** | **$1.41 \times 10^{-2}$** | **pink** |
| ASCC1 | Q8N9N2 | -0.38 | $1.52 \times 10^{-2}$ | turquoise |
| IFI30 | P13284 | 0.26 | $1.52 \times 10^{-2}$ | turquoise |
| OMG | P23515 | -0.62 | $1.52 \times 10^{-2}$ | purple |
| TNFAIP6 | P98066 | 0.38 | $1.52 \times 10^{-2}$ | grey |
| CC2D1A | Q6P1N0 | -0.25 | $1.57 \times 10^{-2}$ | yellow |
| EIF4G1 | Q04637 | -0.38 | $1.61 \times 10^{-2}$ | turquoise |
| NIT1 | Q86X76 | -0.26 | $1.61 \times 10^{-2}$ | yellow |
| BRAP | Q7Z569 | -0.43 | $1.68 \times 10^{-2}$ | turquoise |
| **PALM3** | **A6NDB9** | **0.58** | **$1.68 \times 10^{-2}$** | **turquoise** |
| DNAJB1 | P25685 | -0.42 | $1.73 \times 10^{-2}$ | yellow |
| PDGFRA | P16234 | 0.18 | $1.79 \times 10^{-2}$ | turquoise |
| THBS4 | P35443 | 0.32 | $1.79 \times 10^{-2}$ | magenta |
| BACH1 | O14867 | -0.28 | $1.86 \times 10^{-2}$ | turquoise |
| ARHGAP10 | A1A4S6 | -0.49 | $1.88 \times 10^{-2}$ | turquoise |
| CNP | P09543 | -0.30 | $1.88 \times 10^{-2}$ | yellow |
| PTGES2 | Q9H7Z7 | 0.37 | $1.89 \times 10^{-2}$ | magenta |
| APRT | P07741 | -0.27 | $1.91 \times 10^{-2}$ | turquoise |
| DNAJB4 | Q9UDY4 | 0.23 | $1.91 \times 10^{-2}$ | magenta |
| NCAM1 | P13591 | 0.17 | $1.91 \times 10^{-2}$ | grey |
| NDUFB7 | P17568 | -0.28 | $1.91 \times 10^{-2}$ | turquoise |
| SIGLEC8 | Q9NYZ4 | 0.30 | $1.91 \times 10^{-2}$ | turquoise |
| TNIP1 | Q15025 | -0.37 | $1.91 \times 10^{-2}$ | black |
| VPS37A | Q8NEZ2 | -0.35 | $1.91 \times 10^{-2}$ | turquoise |

| Protein | UniProt ID | log2 fold change | FDR-adjusted P value | WGCNA module |
|---|---|---|---|---|
| PTH | P01270 | -0.42 | $1.93 \times 10^{-2}$ | grey |
| LPP | Q93052 | -0.29 | $1.96 \times 10^{-2}$ | turquoise |
| SEC31A | O94979 | -0.34 | $1.96 \times 10^{-2}$ | turquoise |
| CD38 | P28907 | -0.20 | $2.00 \times 10^{-2}$ | yellow |
| SCN4B | Q8IWT1 | -0.25 | $2.00 \times 10^{-2}$ | brown |
| SARG | Q9BW04 | -0.42 | $2.01 \times 10^{-2}$ | turquoise |
| CEP250 | Q9BV73 | -0.47 | $2.09 \times 10^{-2}$ | black |
| DCTN1 | Q14203 | -0.33 | $2.09 \times 10^{-2}$ | yellow |
| DDHD2 | O94830 | -0.23 | $2.09 \times 10^{-2}$ | turquoise |
| FLT3LG | P49771 | -0.21 | $2.09 \times 10^{-2}$ | grey |
| GIT1 | Q9Y2X7 | -0.32 | $2.09 \times 10^{-2}$ | black |
| TGFBR1 | P36897 | 0.34 | $2.09 \times 10^{-2}$ | yellow |
| C2orf88 | Q9BSF0 | -0.38 | $2.12 \times 10^{-2}$ | turquoise |
| CLEC4G | Q6UXB4 | 0.19 | $2.12 \times 10^{-2}$ | turquoise |
| ITGAV | P06756 | -0.15 | $2.12 \times 10^{-2}$ | grey |
| PLEKHO1 | Q53GL0 | -0.42 | $2.12 \times 10^{-2}$ | turquoise |
| **TMPRSS15** | **P98073** | **-0.54** | $\mathbf{2.12 \times 10^{-2}}$ | **grey** |
| CDKN1A | P38936 | -0.39 | $2.17 \times 10^{-2}$ | turquoise |
| LIMD1 | Q9UGP4 | -0.33 | $2.19 \times 10^{-2}$ | turquoise |
| EBAG9 | O00559 | -0.35 | $2.22 \times 10^{-2}$ | turquoise |
| PDLIM5 | Q96HC4 | -0.44 | $2.26 \times 10^{-2}$ | turquoise |
| PLXNA4 | Q9HCM2 | -0.26 | $2.26 \times 10^{-2}$ | turquoise |
| BOC | Q9BWV1 | 0.13 | $2.31 \times 10^{-2}$ | grey |
| CDKN2D | P55273 | -0.49 | $2.31 \times 10^{-2}$ | turquoise |
| EPS8L2 | Q9H6S3 | -0.18 | $2.31 \times 10^{-2}$ | turquoise |
| HCLS1 | P14317 | -0.28 | $2.31 \times 10^{-2}$ | turquoise |
| IL17RB | Q9NRM6 | 0.29 | $2.31 \times 10^{-2}$ | turquoise |
| MDK | P21741 | 0.29 | $2.31 \times 10^{-2}$ | turquoise |
| RANBP1 | P43487 | -0.29 | $2.31 \times 10^{-2}$ | yellow |
| SDCCAG8 | Q86SQ7 | -0.40 | $2.31 \times 10^{-2}$ | black |
| **TEX35** | **Q5T0J7** | **0.72** | $\mathbf{2.31 \times 10^{-2}}$ | **grey** |

| Protein | UniProt ID | log2 fold change | FDR-adjusted P value | WGCNA module |
|---|---|---|---|---|
| NFKB1 | P19838 | -0.34 | $2.38 \times 10^{-2}$ | turquoise |
| PTX3 | P26022 | 0.33 | $2.39 \times 10^{-2}$ | turquoise |
| GOLGA3 | Q08378 | -0.39 | $2.56 \times 10^{-2}$ | yellow |
| MSTN | O14793 | -0.30 | $2.57 \times 10^{-2}$ | grey |
| CCL21 | O00585 | 0.28 | $2.63 \times 10^{-2}$ | turquoise |
| GSN | P06396 | -0.12 | $2.63 \times 10^{-2}$ | brown |
| **STAT5B** | **P51692** | **-0.54** | **$2.63 \times 10^{-2}$** | **turquoise** |
| ICA1 | Q05084 | -0.28 | $2.68 \times 10^{-2}$ | turquoise |
| FGL1 | Q08830 | 0.39 | $2.71 \times 10^{-2}$ | turquoise |
| VEGFB | P49765 | 0.19 | $2.71 \times 10^{-2}$ | turquoise |
| **YARS1** | **P54577** | **-0.60** | **$2.71 \times 10^{-2}$** | **turquoise** |
| ASAH2 | Q9NR71 | -0.26 | $2.80 \times 10^{-2}$ | grey |
| CHCHD6 | Q9BRQ6 | 0.20 | $2.80 \times 10^{-2}$ | turquoise |
| SNAP23 | O00161 | -0.40 | $2.80 \times 10^{-2}$ | turquoise |
| ADD1 | P35611 | -0.32 | $2.88 \times 10^{-2}$ | red |
| MTSS2 | Q765P7 | -0.42 | $2.88 \times 10^{-2}$ | turquoise |
| PIBF1 | Q8WXW3 | -0.36 | $2.88 \times 10^{-2}$ | turquoise |
| PALM2 | Q8IXS6 | 0.25 | $2.97 \times 10^{-2}$ | turquoise |
| BMERB1 | Q96MC5 | -0.20 | $3.04 \times 10^{-2}$ | grey |
| TNFRSF9 | Q07011 | 0.27 | $3.04 \times 10^{-2}$ | turquoise |
| GFAP | P14136 | 0.31 | $3.07 \times 10^{-2}$ | purple |
| LRCH4 | O75427 | -0.25 | $3.07 \times 10^{-2}$ | blue |
| RABEP1 | Q15276 | -0.23 | $3.07 \times 10^{-2}$ | yellow |
| TXLNA | P40222 | -0.29 | $3.07 \times 10^{-2}$ | turquoise |
| ATE1 | O95260 | -0.40 | $3.08 \times 10^{-2}$ | turquoise |
| PRDX5 | P30044 | -0.32 | $3.22 \times 10^{-2}$ | turquoise |
| BCHE | P06276 | -0.14 | $3.25 \times 10^{-2}$ | grey |
| **MAMDC4** | **Q6UXC1** | **-0.58** | **$3.29 \times 10^{-2}$** | **green** |
| LCP2 | Q13094 | -0.47 | $3.31 \times 10^{-2}$ | turquoise |
| USO1 | O60763 | -0.35 | $3.31 \times 10^{-2}$ | turquoise |
| CAP2 | P40123 | 0.45 | $3.36 \times 10^{-2}$ | magenta |

| Protein | UniProt ID | log2 fold change | FDR-adjusted P value | WGCNA module |
|---|---|---|---|---|
| OTUD7B | Q6GQQ9 | -0.20 | $3.36 \times 10^{-2}$ | turquoise |
| DNAJA2 | O60884 | -0.26 | $3.37 \times 10^{-2}$ | turquoise |
| FGF21 | Q9NSA1 | 0.98 | $3.37 \times 10^{-2}$ | turquoise |
| **PSG1** | **P11464** | **0.72** | **$3.37 \times 10^{-2}$** | **turquoise** |
| CXADR | P78310 | 0.29 | $3.38 \times 10^{-2}$ | turquoise |
| GMPR | P36959 | -0.30 | $3.38 \times 10^{-2}$ | yellow |
| OTUD6B | Q8N6M0 | -0.32 | $3.38 \times 10^{-2}$ | turquoise |
| SLC39A14 | Q15043 | 0.19 | $3.38 \times 10^{-2}$ | turquoise |
| CTSL | P07711 | 0.19 | $3.49 \times 10^{-2}$ | turquoise |
| ESM1 | Q9NQ30 | 0.20 | $3.49 \times 10^{-2}$ | turquoise |
| CRTAM | O95727 | 0.28 | $3.61 \times 10^{-2}$ | turquoise |
| LRRN1 | Q6UXK5 | 0.22 | $3.61 \times 10^{-2}$ | magenta |
| LAG3 | P18627 | 0.24 | $3.62 \times 10^{-2}$ | turquoise |
| PDCD1LG2 | Q9BQ51 | 0.16 | $3.63 \times 10^{-2}$ | turquoise |
| CDK17 | Q00537 | -0.40 | $3.67 \times 10^{-2}$ | turquoise |
| CEND1 | Q8N111 | 0.37 | $3.67 \times 10^{-2}$ | grey |
| CXCL8 | P10145 | 0.28 | $3.67 \times 10^{-2}$ | pink |
| IL6 | P05231 | 0.48 | $3.67 \times 10^{-2}$ | turquoise |
| PDE2A | O00408 | -0.37 | $3.67 \times 10^{-2}$ | turquoise |
| CSF1 | P09603 | 0.15 | $3.68 \times 10^{-2}$ | brown |
| LRBA | P50851 | -0.39 | $3.68 \times 10^{-2}$ | turquoise |
| GFRA1 | P56159 | 0.18 | $3.85 \times 10^{-2}$ | turquoise |
| EVI5 | O60447 | -0.26 | $4.02 \times 10^{-2}$ | turquoise |
| DNAJC6 | O75061 | -0.30 | $4.03 \times 10^{-2}$ | yellow |
| TNFRSF13B | O14836 | 0.19 | $4.05 \times 10^{-2}$ | turquoise |
| GPC2 | Q8N158 | 0.19 | $4.08 \times 10^{-2}$ | magenta |
| SKAP2 | O75563 | -0.41 | $4.08 \times 10^{-2}$ | turquoise |
| BOLA2 | Q9H3K6 | -0.23 | $4.10 \times 10^{-2}$ | red |
| DLL1 | O00548 | 0.16 | $4.10 \times 10^{-2}$ | brown |
| FGFBP3 | Q8TAT2 | 0.20 | $4.10 \times 10^{-2}$ | turquoise |
| IL12A_IL12B | P29459_P29460 | 0.49 | $4.10 \times 10^{-2}$ | yellow |

| Protein | UniProt ID | log2 fold change | FDR-adjusted P value | WGCNA module |
|---|---|---|---|---|
| IRAK4 | Q9NWZ3 | -0.39 | $4.10 \times 10^{-2}$ | turquoise |
| NT5C3A | Q9H0P0 | -0.38 | $4.10 \times 10^{-2}$ | turquoise |
| RGS6 | P49758 | -0.52 | $4.10 \times 10^{-2}$ | turquoise |
| **SAMD14** | **Q8IZD0** | **-0.50** | **$4.10 \times 10^{-2}$** | **turquoise** |
| SIGLEC10 | Q96LC7 | 0.18 | $4.10 \times 10^{-2}$ | turquoise |
| TMED8 | Q6PL24 | -0.41 | $4.10 \times 10^{-2}$ | turquoise |
| TNFRSF4 | P43489 | 0.25 | $4.10 \times 10^{-2}$ | turquoise |
| IST1 | P53990 | -0.33 | $4.17 \times 10^{-2}$ | turquoise |
| **ANKRD11** | **Q6UB99** | **-0.79** | **$4.19 \times 10^{-2}$** | **grey** |
| ACTA2 | P62736 | 0.19 | $4.21 \times 10^{-2}$ | turquoise |
| TYMS | P04818 | 0.33 | $4.21 \times 10^{-2}$ | turquoise |
| ANGPTL4 | Q9BY76 | 0.19 | $4.21 \times 10^{-2}$ | turquoise |
| EBI3_IL27 | Q14213_Q8NEV9 | 0.18 | $4.21 \times 10^{-2}$ | turquoise |
| FOLR2 | P14207 | 0.16 | $4.21 \times 10^{-2}$ | turquoise |
| SPAG9 | O60271 | -0.23 | $4.21 \times 10^{-2}$ | turquoise |
| YES1 | P07947 | -0.28 | $4.21 \times 10^{-2}$ | turquoise |
| NAA80 | Q93015 | -0.42 | $4.23 \times 10^{-2}$ | turquoise |
| STAMBP | O95630 | -0.23 | $4.23 \times 10^{-2}$ | yellow |
| LDLRAP1 | Q5SW96 | -0.45 | $4.24 \times 10^{-2}$ | turquoise |
| **MYL6B** | **P14649** | **0.99** | **$4.24 \times 10^{-2}$** | **grey** |
| ISM1 | B1AKI9 | -0.20 | $4.29 \times 10^{-2}$ | yellow |
| CRKL | P46109 | -0.31 | $4.29 \times 10^{-2}$ | turquoise |
| PTPRK | Q15262 | 0.16 | $4.29 \times 10^{-2}$ | turquoise |
| CLEC10A | Q8IUN9 | -0.19 | $4.39 \times 10^{-2}$ | yellow |
| BAG5 | Q9UL15 | -0.30 | $4.43 \times 10^{-2}$ | yellow |
| DCTN2 | Q13561 | -0.30 | $4.43 \times 10^{-2}$ | turquoise |
| PPP1R18 | Q6NYC8 | -0.20 | $4.43 \times 10^{-2}$ | blue |
| TADA3 | O75528 | -0.34 | $4.43 \times 10^{-2}$ | turquoise |
| MRC1 | P22897 | 0.17 | $4.45 \times 10^{-2}$ | turquoise |
| RILP | Q96NA2 | -0.25 | $4.45 \times 10^{-2}$ | red |
| RRM2 | P31350 | 0.32 | $4.45 \times 10^{-2}$ | turquoise |

| Protein | UniProt ID | log2 fold change | FDR-adjusted P value | WGCNA module |
|---|---|---|---|---|
| SLIT2 | O94813 | 0.21 | $4.45 \times 10^{-2}$ | turquoise |
| CEACAM8 | P31997 | 0.26 | $4.49 \times 10^{-2}$ | blue |
| SIRT2 | Q8IXJ6 | -0.28 | $4.49 \times 10^{-2}$ | yellow |
| TACC3 | Q9Y6A5 | -0.44 | $4.49 \times 10^{-2}$ | turquoise |
| GOPC | Q9HD26 | -0.41 | $4.54 \times 10^{-2}$ | turquoise |
| CDC37 | Q16543 | -0.32 | $4.55 \times 10^{-2}$ | turquoise |
| HLA-A | P04439 | 0.28 | $4.58 \times 10^{-2}$ | yellow |
| RAB11FIP3 | O75154 | -0.44 | $4.58 \times 10^{-2}$ | turquoise |
| TGFA | P01135 | 0.15 | $4.58 \times 10^{-2}$ | turquoise |
| UMAD1 | C9J7I0 | -0.26 | $4.58 \times 10^{-2}$ | yellow |
| NPY | P01303 | -0.36 | $4.60 \times 10^{-2}$ | grey |
| SERPINB9 | P50453 | -0.15 | $4.61 \times 10^{-2}$ | turquoise |
| CBS | P35520 | 0.44 | $4.67 \times 10^{-2}$ | green |
| MMP8 | P22894 | 0.40 | $4.70 \times 10^{-2}$ | blue |
| **MYH7** | **P12883** | **0.62** | **$4.70 \times 10^{-2}$** | **magenta** |
| LEPR | P48357 | 0.17 | $4.84 \times 10^{-2}$ | turquoise |
| **DOK1** | **Q99704** | **-0.51** | **$4.90 \times 10^{-2}$** | **turquoise** |

Abbreviations: FDR: false discovery rate; WGCNA: weighted gene co-expression network analysis.

**eTable 2. Module colors ALS versus control.** Module colors and number of proteins in each module.

| black | Blue | Brown | Green | Grey | Magenta | Pink | Purple | Red | Turquoise | yellow |
|---|---|---|---|---|---|---|---|---|---|---|
| 60 | 184 | 136 | 96 | 553 | 51 | 57 | 37 | 92 | 1012 | 133 |

**eTable 3. Hub proteins per co-expression module.** Hub proteins were defined as proteins with absolute module membership > 0.80 among the top 10 proteins per module. Module membership (MM) reflects the Pearson correlation between protein expression and the module eigengene. All except purple, brown and pink modules had 10 hub proteins meeting the criteria.

| Gene | WGCNA Module | MM |
|---|---|---:|
| TRIP11 | black | 0.9332107 |
| RAB3IP | black | 0.8868019 |
| KIFC3 | black | 0.8834437 |
| TMF1 | black | 0.8771452 |
| RABEP2 | black | 0.8744886 |
| SNX29 | black | 0.8683233 |
| SDCCAG8 | black | 0.8567372 |
| CALCOCO1 | black | 0.8524764 |
| CAMSAP1 | black | 0.8521615 |
| CEP131 | black | 0.8507973 |
| SMNDC1 | blue | 0.9204250 |
| TCERG1 | blue | 0.9132133 |
| ELOA | blue | 0.9086622 |
| MORC3 | blue | 0.9048843 |
| BAP18 | blue | 0.9037535 |
| SNRPB2 | blue | 0.8920153 |
| WDR46 | blue | 0.8803379 |
| C7orf50 | blue | 0.8791014 |
| ARHGAP27 | blue | 0.8767259 |
| ANP32E | blue | 0.8748891 |
| TGFBR2 | brown | 0.8743095 |
| JAM2 | brown | 0.8524539 |
| NBL1 | brown | 0.8400590 |
| NECTIN4 | brown | 0.8196370 |
| RGMB | brown | 0.8186062 |
| BTN2A1 | brown | 0.8076272 |
| PCBD1 | green | 0.9017285 |
| ACY1 | green | 0.8909653 |

| | | |
|---|---|---|
| FTCD | green | 0.8718484 |
| DCXR | green | 0.8467682 |
| SPATA2L | green | 0.8464343 |
| ADH1B | green | 0.8437805 |
| AGXT | green | 0.8426124 |
| RIDA | green | 0.8354883 |
| KRT18 | green | 0.8219373 |
| ADH4 | green | 0.8193822 |
| MYBPC1 | magenta | 0.9305012 |
| HSPB6 | magenta | 0.9148327 |
| CSRP3 | magenta | 0.9045913 |
| CMYA5 | magenta | 0.8957576 |
| DTNB | magenta | 0.8923062 |
| ACTN2 | magenta | 0.8916224 |
| CORO6 | magenta | 0.8727299 |
| MYBPH | magenta | 0.8690712 |
| TNNC1 | magenta | 0.8562167 |
| MYL3 | magenta | 0.8462427 |
| SPARC | pink | 0.9212256 |
| PDGFB | pink | 0.9150593 |
| ANGPT1 | pink | 0.9074270 |
| VEGFC | pink | 0.9061562 |
| PDGFD | pink | 0.8990744 |
| DKK1 | pink | 0.8857119 |
| CPXM1 | pink | 0.8449401 |
| CCN2 | pink | 0.8371805 |
| SERPINE2 | pink | 0.8189743 |
| PTPRR | purple | 0.8444860 |
| MOG | purple | 0.8383320 |
| PTPRN | purple | 0.8303703 |
| BCAN | purple | 0.8272030 |

| | | |
|---|---|---|
| PTPRN2 | purple | 0.8253364 |
| PSMG3 | red | 0.9396583 |
| ADSL | red | 0.9071074 |
| HAGH | red | 0.9069976 |
| YOD1 | red | 0.8861772 |
| DNPH1 | red | 0.8858307 |
| PRDX2 | red | 0.8776479 |
| SH3GLB2 | red | 0.8742371 |
| C9orf40 | red | 0.8726886 |
| AARSD1 | red | 0.8598475 |
| CA1 | red | 0.8578732 |
| ASCC1 | turquoise | 0.9193700 |
| PLA2G4A | turquoise | 0.9079615 |
| CRKL | turquoise | 0.9070953 |
| DBNL | turquoise | 0.9066599 |
| CFAP36 | turquoise | 0.9032076 |
| GRAP2 | turquoise | 0.8965030 |
| PLCB2 | turquoise | 0.8950365 |
| CACYBP | turquoise | 0.8946138 |
| PYM1 | turquoise | 0.8935702 |
| TBCC | turquoise | 0.8929853 |
| PLPBP | yellow | 0.9077158 |
| RWDD1 | yellow | 0.8962847 |
| DTYMK | yellow | 0.8907045 |
| TBCA | yellow | 0.8819356 |
| GGCT | yellow | 0.8807158 |
| GMPR2 | yellow | 0.8806453 |
| CIAPIN1 | yellow | 0.8693206 |
| IMPACT | yellow | 0.8672934 |
| IGBP1 | yellow | 0.8621152 |
| SGTA | yellow | 0.8601767 |

Abbreviations: MM: module membership; WGCNA: weighted gene co-expression network analysis.

**eTable 4. Module-trait association ALS versus control from weighted gene co-expression network analysis.** Pearson correlation (r) coefficient and p-values (p). Significant ($p<0.05$) are in bold.

| WGCNA Module | Trait | Pearson r | P value |
|---|---|---|---|
| **magenta** | **ALS** | **0.419** | **$3.1 \times 10^{-16}$** |
| **turquoise** | **ALS** | **-0.155** | **$3.6 \times 10^{-3}$** |
| **yellow** | **ALS** | **-0.142** | **$7.7 \times 10^{-3}$** |
| red | ALS | -0.102 | $5.7 \times 10^{-2}$ |
| black | ALS | -0.101 | $6.0 \times 10^{-2}$ |
| pink | ALS | 0.099 | $6.5 \times 10^{-2}$ |
| green | ALS | 0.067 | $2.1 \times 10^{-1}$ |
| purple | ALS | -0.066 | $2.2 \times 10^{-1}$ |
| brown | ALS | -0.010 | $8.6 \times 10^{-1}$ |
| blue | ALS | 0.005 | $9.3 \times 10^{-1}$ |
| **magenta** | **Control** | **-0.419** | **$3.1 \times 10^{-16}$** |
| **turquoise** | **Control** | **0.155** | **$3.6 \times 10^{-3}$** |
| **yellow** | **Control** | **0.142** | **$7.7 \times 10^{-3}$** |
| red | Control | 0.102 | $5.7 \times 10^{-2}$ |
| black | Control | 0.101 | $6.0 \times 10^{-2}$ |
| pink | Control | -0.099 | $6.5 \times 10^{-2}$ |
| green | Control | -0.067 | $2.1 \times 10^{-1}$ |
| purple | Control | 0.066 | $2.2 \times 10^{-1}$ |
| brown | Control | 0.010 | $8.6 \times 10^{-1}$ |
| blue | Control | -0.005 | $9.3 \times 10^{-1}$ |

Abbreviations: ALS: Amyotrophic Lateral Sclerosis; WGCNA: weighted gene co-expression network analysis.

**eTable 5. Logistic regression ALS versus controls.** All hub proteins from modules significantly associated with ALS patients (n=299) were included and compared to controls (n=50) in a logistic regression. Except for the covariates adjusted normalized protein expression, no additional covariates were included in the model. P values are FDR-adjusted (Benjamini–Hochberg). Proteins are ordered by ascending FDR-adjusted P value. Significant proteins (FDR < 0.05 and AUC > 0.65) are in bold.

| Protein | UniProt ID | OR (95% CI) | AUC | FDR-adjusted P value | WGCNA Module |
|---|---|---|---|---|---|
| **MYBPC1** | **Q00872** | **3.07 (2.27–4.31)** | **0.842** | **$6.63 \times 10^{-11}$** | **magenta** |
| **CSRP3** | **P50461** | **2.58 (2.01–3.44)** | **0.862** | **$6.63 \times 10^{-11}$** | **magenta** |
| **ACTN2** | **P35609** | **3.02 (2.22–4.25)** | **0.825** | **$1.87 \times 10^{-10}$** | **magenta** |
| **MYBPH** | **Q13203** | **2.5 (1.93–3.34)** | **0.838** | **$3.94 \times 10^{-10}$** | **magenta** |
| **CORO6** | **Q6QEF8** | **5.64 (3.48–9.9)** | **0.864** | **$4.17 \times 10^{-10}$** | **magenta** |
| **HSPB6** | **O14558** | **11.05 (5.49–24.25)** | **0.821** | **$9.55 \times 10^{-10}$** | **magenta** |
| **DTNB** | **O60941** | **12.28 (5.86–28.19)** | **0.815** | **$1.33 \times 10^{-9}$** | **magenta** |
| **TNNC1** | **P63316** | **2.03 (1.65–2.57)** | **0.785** | **$1.33 \times 10^{-9}$** | **magenta** |
| **CMYA5** | **Q8N3K9** | **2.65 (1.97–3.67)** | **0.792** | **$2.44 \times 10^{-9}$** | **magenta** |
| **MYL3** | **P08590** | **2.39 (1.78–3.29)** | **0.762** | **$5.73 \times 10^{-8}$** | **magenta** |
| **IMPACT** | **Q9P2X3** | **0.34 (0.18–0.63)** | **0.656** | **$1.74 \times 10^{-3}$** | **yellow** |
| **ASCC1** | **Q8N9N2** | **0.46 (0.29–0.72)** | **0.653** | **$2.19 \times 10^{-3}$** | **turquoise** |
| CRKL | P46109 | 0.51 (0.32–0.81) | 0.622 | $1.11 \times 10^{-2}$ | turquoise |
| DTYMK | P23919 | 0.47 (0.27–0.81) | 0.627 | $1.45 \times 10^{-2}$ | yellow |
| PLCB2 | Q00722 | 0.68 (0.5–0.9) | 0.611 | $1.92 \times 10^{-2}$ | turquoise |
| GMPR2 | Q9P2T1 | 0.5 (0.29–0.85) | 0.620 | $2.16 \times 10^{-2}$ | yellow |
| DBNL | Q9UJU6 | 0.59 (0.39–0.89) | 0.608 | $2.25 \times 10^{-2}$ | turquoise |
| CACYBP | Q9HB71 | 0.63 (0.43–0.9) | 0.604 | $2.25 \times 10^{-2}$ | turquoise |
| CIAPIN1 | Q6FI81 | 0.45 (0.23–0.86) | 0.598 | $2.83 \times 10^{-2}$ | yellow |
| PYM1 | Q9BRP8 | 0.7 (0.51–0.94) | 0.593 | $2.89 \times 10^{-2}$ | turquoise |
| SGTA | O43765 | 0.51 (0.29–0.9) | 0.603 | $2.89 \times 10^{-2}$ | yellow |
| TBCC | Q15814 | 0.56 (0.34–0.92) | 0.605 | $3.01 \times 10^{-2}$ | turquoise |
| RWDD1 | Q9H446 | 0.48 (0.25–0.89) | 0.590 | $3.01 \times 10^{-2}$ | yellow |
| TBCA | O75347 | 0.61 (0.39–0.93) | 0.603 | $3.01 \times 10^{-2}$ | yellow |
| GRAP2 | O75791 | 0.71 (0.51–0.95) | 0.594 | $3.32 \times 10^{-2}$ | turquoise |
| GGCT | O75223 | 0.51 (0.27–0.95) | 0.586 | $4.10 \times 10^{-2}$ | yellow |
| IGBP1 | P78318 | 0.59 (0.35–0.96) | 0.588 | $4.10 \times 10^{-2}$ | yellow |

| Protein | UniProt ID | OR (95% CI) | AUC | FDR-adjusted P value | WGCNA Module |
|---|---|---|---|---|---|
| PLA2G4A | P47712 | 0.62 (0.39–0.97) | 0.577 | $4.43 \times 10^{-2}$ | turquoise |
| CFAP36 | Q96G28 | 0.67 (0.41–1.07) | 0.576 | $1.01 \times 10^{-1}$ | turquoise |
| PLPBP | O94903 | 0.64 (0.36–1.11) | 0.562 | $1.19 \times 10^{-1}$ | yellow |

Abbreviations: AUC: area under the curve; CI: confidence interval; FDR: false discovery rate; OR: odds ratio; WGCNA: weighted gene co-expression network analysis.

**eTable 6. Differentially expressed proteins Spinal versus Bulbar onset.** Models were adjusted for ALSFRS-R score and BMI. P values are FDR-adjusted (Benjamini–Hochberg). All proteins meeting FDR < 0.05 are displayed. Proteins meeting criteria FDR <0.05 and |log2 fold change| > 0.5 are in bold. P values are FDR-adjusted (Benjamini–Hochberg). Ordered with ascending FDR-adjusted p-value.

| Protein | UniProt ID | log2 fold change | FDR-adjusted P value | WGCNA module |
|---|---|---|---|---|
| EDA2R | Q9HAV5 | 0.54 | $4.40 \times 10^{-4}$ | magenta |
| MEGF10 | Q96KG7 | 0.41 | $6.26 \times 10^{-4}$ | magenta |
| CORO6 | Q6QEF8 | 0.74 | $6.88 \times 10^{-4}$ | magenta |
| DTNB | O60941 | 0.35 | $6.94 \times 10^{-4}$ | magenta |
| HSPB6 | O14558 | 0.36 | $1.03 \times 10^{-3}$ | magenta |
| AFAP1L1 | Q8TED9 | 0.29 | $1.44 \times 10^{-3}$ | magenta |
| HPCAL4 | Q9UM19 | 0.41 | $5.84 \times 10^{-3}$ | magenta |
| ART3 | Q13508 | -0.28 | $6.42 \times 10^{-3}$ | brown |
| MYBPC1 | Q00872 | 0.67 | $6.42 \times 10^{-3}$ | magenta |
| MYBPH | Q13203 | 0.87 | $6.42 \times 10^{-3}$ | magenta |
| SLMAP | Q14BN4 | 0.35 | $6.42 \times 10^{-3}$ | magenta |
| CHCHD10 | Q8WYQ3 | 0.28 | $6.42 \times 10^{-3}$ | magenta |
| ADAM12 | O43184 | 0.51 | $7.96 \times 10^{-3}$ | turquoise |
| RBFOX3 | A6NFN3 | 0.37 | $8.65 \times 10^{-3}$ | magenta |
| CSRP3 | P50461 | 0.91 | $8.71 \times 10^{-3}$ | magenta |
| SORBS1 | Q9BX66 | 0.36 | $8.99 \times 10^{-3}$ | magenta |
| DNAJB4 | Q9UDY4 | 0.24 | $9.21 \times 10^{-3}$ | magenta |
| DMD | P11532 | 0.30 | $9.83 \times 10^{-3}$ | magenta |
| MYOM3 | Q5VTT5 | 0.63 | $1.22 \times 10^{-2}$ | magenta |
| MYBPC2 | Q14324 | 0.67 | $2.28 \times 10^{-2}$ | magenta |
| USP28 | Q96RU2 | 0.30 | $2.28 \times 10^{-2}$ | magenta |
| LMNB2 | Q03252 | 0.20 | $2.31 \times 10^{-2}$ | turquoise |
| CMYA5 | Q8N3K9 | 0.57 | $3.62 \times 10^{-2}$ | magenta |
| CRNN | Q9UBG3 | -0.49 | $3.62 \times 10^{-2}$ | grey |

Abbreviations: ALSFRS-R: ALS Functional Rating Scale–Revised; BMI: body mass index; FDR: false discovery rate; WGCNA: weighted gene co-expression network analysis.

**eTable 7. Module-trait association ALS-specific traits from weighted gene co-expression network analysis.** ECAS score (n=156), ALSFRS-R score (n=251), spinal onset (n=184), bulbar onset (n=115), ALS-FTSD (n=71), and non-ALS-FTSD (n=90). Pearson correlation coefficient (r) and p-values (p) are shown.

| WGCNA Module | Trait | Pearson r | P value |
|---|---|---:|---:|
| **brown** | **ALS-FTSD** | **0.227** | **$3.8 \times 10^{-3}$** |
| **yellow** | **ALS-FTSD** | **-0.204** | **$9.4 \times 10^{-3}$** |
| **turquoise** | **ALS-FTSD** | **-0.191** | **$1.5 \times 10^{-2}$** |
| black | ALS-FTSD | -0.139 | $8.0 \times 10^{-2}$ |
| red | ALS-FTSD | -0.111 | $1.6 \times 10^{-1}$ |
| magenta | ALS-FTSD | -0.098 | $2.1 \times 10^{-1}$ |
| green | ALS-FTSD | 0.051 | $5.2 \times 10^{-1}$ |
| pink | ALS-FTSD | 0.051 | $5.2 \times 10^{-1}$ |
| purple | ALS-FTSD | 0.035 | $6.6 \times 10^{-1}$ |
| blue | ALS-FTSD | -0.019 | $8.1 \times 10^{-1}$ |
| **turquoise** | **ALSFRS-R score** | **0.216** | **$5.8 \times 10^{-4}$** |
| **black** | **ALSFRS-R score** | **0.150** | **$1.8 \times 10^{-2}$** |
| **magenta** | **ALSFRS-R score** | **-0.126** | **$4.6 \times 10^{-2}$** |
| brown | ALSFRS-R score | -0.094 | $1.4 \times 10^{-1}$ |
| pink | ALSFRS-R score | -0.076 | $2.3 \times 10^{-1}$ |
| yellow | ALSFRS-R score | 0.073 | $2.5 \times 10^{-1}$ |
| red | ALSFRS-R score | -0.061 | $3.4 \times 10^{-1}$ |
| purple | ALSFRS-R score | -0.050 | $4.3 \times 10^{-1}$ |
| blue | ALSFRS-R score | -0.024 | $7.1 \times 10^{-1}$ |
| green | ALSFRS-R score | -0.007 | $9.1 \times 10^{-1}$ |
| **magenta** | **Bulbar** | **-0.320** | **$1.6 \times 10^{-8}$** |
| blue | Bulbar | -0.058 | $3.2 \times 10^{-1}$ |
| brown | Bulbar | -0.050 | $3.9 \times 10^{-1}$ |
| pink | Bulbar | -0.042 | $4.7 \times 10^{-1}$ |
| turquoise | Bulbar | 0.038 | $5.2 \times 10^{-1}$ |
| black | Bulbar | -0.015 | $8.0 \times 10^{-1}$ |
| purple | Bulbar | 0.014 | $8.1 \times 10^{-1}$ |
| yellow | Bulbar | 0.011 | $8.5 \times 10^{-1}$ |

| WGCNA Module | Trait | Pearson r | P value |
|---|---|---:|---:|
| red | Bulbar | 0.001 | $9.8 \times 10^{-1}$ |
| green | Bulbar | 0.001 | $9.9 \times 10^{-1}$ |
| brown | ECAS score | -0.148 | $6.5 \times 10^{-2}$ |
| turquoise | ECAS score | 0.100 | $2.1 \times 10^{-1}$ |
| yellow | ECAS score | 0.093 | $2.5 \times 10^{-1}$ |
| magenta | ECAS score | 0.082 | $3.1 \times 10^{-1}$ |
| black | ECAS score | 0.072 | $3.7 \times 10^{-1}$ |
| green | ECAS score | -0.058 | $4.7 \times 10^{-1}$ |
| red | ECAS score | 0.056 | $4.9 \times 10^{-1}$ |
| pink | ECAS score | -0.037 | $6.5 \times 10^{-1}$ |
| purple | ECAS score | -0.031 | $7.0 \times 10^{-1}$ |
| blue | ECAS score | -0.002 | $9.8 \times 10^{-1}$ |
| **purple** | **NEFL** | **0.350** | **$4.9 \times 10^{-10}$** |
| **turquoise** | **NEFL** | **-0.236** | **$3.8 \times 10^{-5}$** |
| **brown** | **NEFL** | **0.207** | **$3.2 \times 10^{-4}$** |
| **yellow** | **NEFL** | **-0.184** | **$1.4 \times 10^{-3}$** |
| **black** | **NEFL** | **-0.163** | **$4.6 \times 10^{-3}$** |
| **magenta** | **NEFL** | **0.139** | **$1.7 \times 10^{-2}$** |
| **blue** | **NEFL** | **-0.115** | **$4.7 \times 10^{-2}$** |
| pink | NEFL | 0.042 | $4.7 \times 10^{-1}$ |
| red | NEFL | -0.021 | $7.2 \times 10^{-1}$ |
| green | NEFL | 0.011 | $8.5 \times 10^{-1}$ |
| **magenta** | **Spinal** | **0.302** | **$9.6 \times 10^{-8}$** |
| blue | Spinal | 0.057 | $3.2 \times 10^{-1}$ |
| brown | Spinal | 0.035 | $5.5 \times 10^{-1}$ |
| pink | Spinal | 0.030 | $6.0 \times 10^{-1}$ |
| turquoise | Spinal | -0.029 | $6.2 \times 10^{-1}$ |
| purple | Spinal | -0.026 | $6.6 \times 10^{-1}$ |
| black | Spinal | 0.020 | $7.3 \times 10^{-1}$ |
| red | Spinal | -0.012 | $8.3 \times 10^{-1}$ |

| WGCNA Module | Trait | Pearson r | P value |
|---|---|---|---|
| yellow | Spinal | -0.012 | $8.4 \times 10^{-1}$ |
| green | Spinal | -0.006 | $9.2 \times 10^{-1}$ |
| **brown** | **non-ALS-FTSD** | **-0.227** | **$3.8 \times 10^{-3}$** |
| **yellow** | **non-ALS-FTSD** | **0.204** | **$9.4 \times 10^{-3}$** |
| **turquoise** | **non-ALS-FTSD** | **0.191** | **$1.5 \times 10^{-2}$** |
| black | non-ALS-FTSD | 0.139 | $8.0 \times 10^{-2}$ |
| red | non-ALS-FTSD | 0.111 | $1.6 \times 10^{-1}$ |
| magenta | non-ALS-FTSD | 0.098 | $2.1 \times 10^{-1}$ |
| green | non-ALS-FTSD | -0.051 | $5.2 \times 10^{-1}$ |
| pink | non-ALS-FTSD | -0.051 | $5.2 \times 10^{-1}$ |
| purple | non-ALS-FTSD | -0.035 | $6.6 \times 10^{-1}$ |
| blue | non-ALS-FTSD | 0.019 | $8.1 \times 10^{-1}$ |

Abbreviations: ALSFRS-R: ALS Functional Rating Scale–Revised; ALS-FTSD: ALS frontotemporal spectrum disorder; ECAS: Edinburgh Cognitive Assessment Scale; WGCNA: weighted gene co-expression network analysis.

**eTable 8. Logistic regression Spinal versus bulbar onset.** Hub proteins from modules significantly associated with site of symptom onset were included. Models were adjusted for ALSFRS-R score and BMI. P values are FDR-adjusted (Benjamini–Hochberg). Significant proteins (FDR < 0.05 and AUC > 0.65) are in bold. Proteins displayed in ascending FDR adjusted p-value.

| Protein | UniProt ID | OR (95% CI) | AUC | FDR-adjusted P value | WGCNA Module |
|---|---|---|---|---|---|
| **DTNB** | **O60941** | **3.66 (2.14–6.51)** | **0.692** | **$2.22 \times 10^{-5}$** | **magenta** |
| **CORO6** | **Q6QEF8** | **1.87 (1.45–2.46)** | **0.685** | **$2.22 \times 10^{-5}$** | **magenta** |
| **HSPB6** | **O14558** | **3.24 (1.97–5.5)** | **0.678** | **$2.26 \times 10^{-5}$** | **magenta** |
| **MYBPC1** | **Q00872** | **1.6 (1.28–2.02)** | **0.656** | **$1.18 \times 10^{-4}$** | **magenta** |
| **MYBPH** | **Q13203** | **1.45 (1.21–1.74)** | **0.664** | **$1.18 \times 10^{-4}$** | **magenta** |
| **CSRP3** | **P50461** | **1.39 (1.19–1.65)** | **0.653** | **$1.29 \times 10^{-4}$** | **magenta** |
| CMYA5 | Q8N3K9 | 1.49 (1.2–1.88) | 0.632 | $7.33 \times 10^{-4}$ | magenta |
| TNNC1 | P63316 | 1.32 (1.11–1.59) | 0.614 | $2.41 \times 10^{-3}$ | magenta |
| ACTN2 | P35609 | 1.45 (1.15–1.85) | 0.622 | $2.61 \times 10^{-3}$ | magenta |
| MYL3 | P08590 | 1.34 (1.06–1.71) | 0.591 | $1.76 \times 10^{-2}$ | magenta |

Abbreviations: ALSFRS-R: ALS Functional Rating Scale–Revised; AUC: area under the curve; BMI: body mass index; CI: confidence interval; FDR: false discovery rate; OR: odds ratio; WGCNA: weighted gene co-expression network analysis.

**eTable 9. Logistic regression: ALS-FTSD vs non-ALS-FTSD.** Hub proteins from modules significantly associated with ALS-FTSD were included. Models were adjusted for site of symptom onset, ALSFRS-R score and BMI. P values are FDR-adjusted (Benjamini–Hochberg). Significant proteins (FDR < 0.05 and AUC > 0.65) are in bold. Proteins displayed in ascending FDR adjusted p-value.

| Protein | UniProt ID | OR (95% CI) | AUC | FDR-adjusted P value | WGCNA Module |
|---|---|---|---|---|---|
| **TBCA** | **O75347** | **0.4 (0.21–0.72)** | **0.741** | **$3.20 \times 10^{-2}$** | **yellow** |
| **ASCC1** | **Q8N9N2** | **0.48 (0.27–0.83)** | **0.740** | **$3.20 \times 10^{-2}$** | **turquoise** |
| **PLA2G4A** | **P47712** | **0.46 (0.25–0.81)** | **0.738** | **$3.20 \times 10^{-2}$** | **turquoise** |
| **JAM2** | **P57087** | **7.48 (2.15–31)** | **0.733** | **$3.20 \times 10^{-2}$** | **brown** |
| **RWDD1** | **Q9H446** | **0.31 (0.13–0.72)** | **0.728** | **$3.20 \times 10^{-2}$** | **yellow** |
| **SGTA** | **O43765** | **0.32 (0.13–0.75)** | **0.728** | **$3.20 \times 10^{-2}$** | **yellow** |
| **CIAPIN1** | **Q6FI81** | **0.32 (0.13–0.73)** | **0.727** | **$3.20 \times 10^{-2}$** | **yellow** |
| **PLPBP** | **O94903** | **0.35 (0.15–0.75)** | **0.724** | **$3.20 \times 10^{-2}$** | **yellow** |
| **DTYMK** | **P23919** | **0.36 (0.16–0.75)** | **0.723** | **$3.20 \times 10^{-2}$** | **yellow** |
| **TBCC** | **Q15814** | **0.44 (0.22–0.84)** | **0.741** | **$3.41 \times 10^{-2}$** | **turquoise** |
| **PLCB2** | **Q00722** | **0.64 (0.44–0.9)** | **0.728** | **$3.41 \times 10^{-2}$** | **turquoise** |
| **NECTIN4** | **Q96NY8** | **3.85 (1.34–12.12)** | **0.718** | **$3.41 \times 10^{-2}$** | **brown** |
| **PYM1** | **Q9BRP8** | **0.64 (0.44–0.92)** | **0.736** | **$3.63 \times 10^{-2}$** | **turquoise** |
| **DBNL** | **Q9UJU6** | **0.57 (0.34–0.91)** | **0.733** | **$3.63 \times 10^{-2}$** | **turquoise** |
| **GMPR2** | **Q9P2T1** | **0.44 (0.21–0.87)** | **0.733** | **$3.63 \times 10^{-2}$** | **yellow** |
| **TGFBR2** | **P37173** | **4.28 (1.32–15.68)** | **0.718** | **$3.63 \times 10^{-2}$** | **brown** |
| **CRKL** | **P46109** | **0.53 (0.3–0.91)** | **0.732** | **$3.67 \times 10^{-2}$** | **turquoise** |
| **GGCT** | **O75223** | **0.38 (0.16–0.88)** | **0.721** | **$4.13 \times 10^{-2}$** | **yellow** |
| **CACYBP** | **Q9HB71** | **0.61 (0.38–0.95)** | **0.725** | **$4.40 \times 10^{-2}$** | **turquoise** |
| **IMPACT** | **Q9P2X3** | **0.36 (0.14–0.9)** | **0.721** | **$4.44 \times 10^{-2}$** | **yellow** |
| CFAP36 | Q96G28 | 0.52 (0.27–0.96) | 0.731 | $5.01 \times 10^{-2}$ | turquoise |
| GRAP2 | O75791 | 0.68 (0.46–0.99) | 0.726 | $5.76 \times 10^{-2}$ | turquoise |
| IGBP1 | P78318 | 0.5 (0.24–1) | 0.725 | $5.90 \times 10^{-2}$ | yellow |
| RGMB | Q6NW40 | 3.4 (1.02–12.3) | 0.715 | $5.90 \times 10^{-2}$ | brown |
| NBL1 | P41271 | 3.48 (1–13.29) | 0.714 | $5.94 \times 10^{-2}$ | brown |
| BTN2A1 | Q7KYR7 | 2.25 (0.83–6.63) | 0.701 | $1.23 \times 10^{-1}$ | brown |

Abbreviations: ALSFRS-R: ALS Functional Rating Scale–Revised; ALS-FTSD: ALS frontotemporal spectrum disorder; AUC: area under the curve; BMI: body mass index; CI: confidence interval; FDR: false discovery rate; OR: odds ratio; WGCNA: weighted gene co-expression network analysis.

**eTable 10. Robust linear regression NEFL.** Hub proteins from modules significantly associated with NEFL expression were included. Models were adjusted for site of symptom onset, ALSFRS-R score, and BMI. Estimates represent the change in NEFL expression per unit increase in covariate-adjusted NPX. P values are FDR-adjusted (Benjamini–Hochberg). Significant proteins (FDR < 0.05) are in bold. Proteins displayed in ascending FDR adjusted p-value.

| Protein | UniProt ID | Estimate (95% CI) | FDR-adjusted P value | WGCNA Module |
|---|---|---|---|---|
| **MOG** | **Q16653** | **0.74 (0.51–0.97)** | **$3.57 \times 10^{-9}$** | **purple** |
| **PTPRR** | **Q15256** | **0.76 (0.52–1)** | **$4.03 \times 10^{-9}$** | **purple** |
| **BCAN** | **Q96GW7** | **0.77 (0.49–1.04)** | **$2.75 \times 10^{-7}$** | **purple** |
| **PTPRN2** | **Q92932** | **0.72 (0.46–0.98)** | **$4.28 \times 10^{-7}$** | **purple** |
| **PTPRN** | **Q16849** | **0.88 (0.55–1.21)** | **$9.86 \times 10^{-7}$** | **purple** |
| **RGMB** | **Q6NW40** | **0.84 (0.5–1.19)** | **$5.68 \times 10^{-6}$** | **brown** |
| **DTYMK** | **P23919** | **-0.37 (-0.58–-0.17)** | **$1.13 \times 10^{-3}$** | **yellow** |
| **IMPACT** | **Q9P2X3** | **-0.42 (-0.66–-0.18)** | **$1.83 \times 10^{-3}$** | **yellow** |
| **NECTIN4** | **Q96NY8** | **0.53 (0.2–0.85)** | **$4.49 \times 10^{-3}$** | **brown** |
| **MYL3** | **P08590** | **0.16 (0.05–0.28)** | **$5.84 \times 10^{-3}$** | **magenta** |
| **ASCC1** | **Q8N9N2** | **-0.3 (-0.49–-0.11)** | **$6.47 \times 10^{-3}$** | **turquoise** |
| **TBCA** | **O75347** | **-0.26 (-0.42–-0.09)** | **$6.66 \times 10^{-3}$** | **yellow** |
| **RWDD1** | **Q9H446** | **-0.37 (-0.62–-0.13)** | **$6.84 \times 10^{-3}$** | **yellow** |
| **CIAPIN1** | **Q6FI81** | **-0.42 (-0.69–-0.14)** | **$8.55 \times 10^{-3}$** | **yellow** |
| **C7orf50** | **Q9BRJ6** | **-0.24 (-0.4–-0.08)** | **$9.11 \times 10^{-3}$** | **blue** |
| **WDR46** | **O15213** | **-0.19 (-0.32–-0.06)** | **$1.02 \times 10^{-2}$** | **blue** |
| **CFAP36** | **Q96G28** | **-0.31 (-0.53–-0.1)** | **$1.20 \times 10^{-2}$** | **turquoise** |
| **JAM2** | **P57087** | **0.49 (0.12–0.85)** | **$1.29 \times 10^{-2}$** | **brown** |
| **IGBP1** | **P78318** | **-0.28 (-0.48–-0.08)** | **$1.42 \times 10^{-2}$** | **yellow** |
| **BAP18** | **Q8IXM2** | **-0.18 (-0.31–-0.05)** | **$1.44 \times 10^{-2}$** | **blue** |
| **TMF1** | **P82094** | **-0.22 (-0.38–-0.06)** | **$1.61 \times 10^{-2}$** | **black** |
| **PLCB2** | **Q00722** | **-0.17 (-0.29–-0.05)** | **$1.61 \times 10^{-2}$** | **turquoise** |
| **BTN2A1** | **Q7KYR7** | **0.47 (0.12–0.81)** | **$1.62 \times 10^{-2}$** | **brown** |
| **CACYBP** | **Q9HB71** | **-0.21 (-0.36–-0.06)** | **$1.65 \times 10^{-2}$** | **turquoise** |
| **ARHGAP27** | **Q6ZUM4** | **-0.29 (-0.52–-0.07)** | **$1.98 \times 10^{-2}$** | **blue** |
| **TBCC** | **Q15814** | **-0.3 (-0.52–-0.08)** | **$2.05 \times 10^{-2}$** | **turquoise** |
| **PLPBP** | **O94903** | **-0.28 (-0.5–-0.05)** | **$2.08 \times 10^{-2}$** | **yellow** |

| Protein | UniProt ID | Estimate (95% CI) | FDR-adjusted P value | WGCNA Module |
|---|---|---|---|---|
| **ACTN2** | **P35609** | **0.14 (0.03–0.26)** | **$2.09 \times 10^{-2}$** | **magenta** |
| **CMYA5** | **Q8N3K9** | **0.13 (0.02–0.23)** | **$2.12 \times 10^{-2}$** | **magenta** |
| **MORC3** | **Q14149** | **-0.2 (-0.36–-0.04)** | **$2.19 \times 10^{-2}$** | **blue** |
| **RAB3IP** | **Q96QF0** | **-0.22 (-0.38–-0.05)** | **$2.26 \times 10^{-2}$** | **black** |
| **CORO6** | **Q6QEF8** | **0.14 (0.03–0.26)** | **$2.36 \times 10^{-2}$** | **magenta** |
| **MYBPC1** | **Q00872** | **0.13 (0.02–0.25)** | **$2.41 \times 10^{-2}$** | **magenta** |
| **SDCCAG8** | **Q86SQ7** | **-0.19 (-0.34–-0.04)** | **$2.44 \times 10^{-2}$** | **black** |
| **CRKL** | **P46109** | **-0.24 (-0.43–-0.06)** | **$2.63 \times 10^{-2}$** | **turquoise** |
| **PYM1** | **Q9BRP8** | **-0.15 (-0.27–-0.03)** | **$2.66 \times 10^{-2}$** | **turquoise** |
| **GGCT** | **O75223** | **-0.3 (-0.55–-0.05)** | **$2.69 \times 10^{-2}$** | **yellow** |
| **PLA2G4A** | **P47712** | **-0.25 (-0.45–-0.05)** | **$2.83 \times 10^{-2}$** | **turquoise** |
| **TRIP11** | **Q15643** | **-0.15 (-0.27–-0.03)** | **$2.95 \times 10^{-2}$** | **black** |
| **MYBPH** | **Q13203** | **0.09 (0.01–0.16)** | **$3.51 \times 10^{-2}$** | **magenta** |
| **DBNL** | **Q9UJU6** | **-0.19 (-0.35–-0.03)** | **$3.73 \times 10^{-2}$** | **turquoise** |
| **GMPR2** | **Q9P2T1** | **-0.25 (-0.47–-0.02)** | **$3.82 \times 10^{-2}$** | **yellow** |
| **CAMSAP1** | **Q5T5Y3** | **-0.17 (-0.32–-0.02)** | **$4.10 \times 10^{-2}$** | **black** |
| **SGTA** | **O43765** | **-0.22 (-0.42–-0.02)** | **$4.46 \times 10^{-2}$** | **yellow** |

Abbreviations: ALSFRS-R: ALS Functional Rating Scale–Revised; BMI: body mass index; CI: confidence interval; FDR: false discovery rate; NEFL: neurofilament light chain; NPX: normalized protein expression; WGCNA: weighted gene co-expression network analysis.

**eTable 11. Robust linear regression ALSFRS-R score and hub protein expression.** Hub proteins from modules significantly associated with ALSFRS-R score were included. Models were adjusted for site of symptom onset and BMI. Estimates represent the change in ALSFRS-R score per unit increase in covariate-adjusted NPX. P values are FDR-adjusted (Benjamini–Hochberg). Significant proteins (FDR < 0.05) are shown in bold.

| Protein | UniProt ID | Estimate (95% CI) | FDR-adjusted P value | WGCNA Module |
|---|---|---|---|---|
| **DTNB** | **O60941** | **-2.59 (-4.13–-1.06)** | **$2.19 \times 10^{-3}$** | **magenta** |
| **RAB3IP** | **Q96QF0** | **1.54 (0.57–2.51)** | **$4.16 \times 10^{-3}$** | **black** |
| **CAMSAP1** | **Q5T5Y3** | **1.48 (0.52–2.44)** | **$5.57 \times 10^{-3}$** | **black** |
| **CEP131** | **Q9UPN4** | **1.16 (0.36–1.95)** | **$9.39 \times 10^{-3}$** | **black** |
| **CORO6** | **Q6QEF8** | **-0.97 (-1.64–-0.29)** | **$1.13 \times 10^{-2}$** | **magenta** |
| **SNX29** | **Q8TEQ0** | **1.02 (0.29–1.74)** | **$1.36 \times 10^{-2}$** | **black** |
| **TRIP11** | **Q15643** | **1.04 (0.26–1.82)** | **$1.86 \times 10^{-2}$** | **black** |
| **TBCC** | **Q15814** | **1.73 (0.42–3.05)** | **$2.09 \times 10^{-2}$** | **turquoise** |
| **PLCB2** | **Q00722** | **0.96 (0.22–1.69)** | **$2.38 \times 10^{-2}$** | **turquoise** |
| **CRKL** | **P46109** | **1.43 (0.32–2.54)** | **$2.44 \times 10^{-2}$** | **turquoise** |
| **CFAP36** | **Q96G28** | **1.87 (0.41–3.34)** | **$2.51 \times 10^{-2}$** | **turquoise** |
| **PLA2G4A** | **P47712** | **1.46 (0.27–2.66)** | **$3.49 \times 10^{-2}$** | **turquoise** |
| **DBNL** | **Q9UJU6** | **1.23 (0.2–2.25)** | **$3.98 \times 10^{-2}$** | **turquoise** |
| **ASCC1** | **Q8N9N2** | **1.27 (0.19–2.34)** | **$4.35 \times 10^{-2}$** | **turquoise** |
| **KIFC3** | **Q9BVG8** | **0.97 (0.14–1.81)** | **$4.58 \times 10^{-2}$** | **black** |

Abbreviations: BMI: body mass index; CI: confidence interval; FDR: false discovery rate; NPX: normalized protein expression; WGCNA: weighted gene co-expression network analysis.
